# Multi-cancer risk stratification based on national health data: A retrospective modelling and validation study

**DOI:** 10.1101/2022.10.12.22280908

**Authors:** Alexander W. Jung, Peter C. Holm, Kumar Gaurav, Jessica Xin Hjaltelin, Davide Placido, Laust Hvas Mortensen, Ewan Birney, Søren Brunak, Moritz Gerstung

## Abstract

**Summary:** *Background:* Health care is experiencing a drive towards digitisation and many countries are implementing national health data resources. Digital medicine promises to identify individuals at elevated risk of disease who may benefit from screening or interventions. This is particularly needed for cancer where early detection improves outcomes. While a range of cancer risk models exists, the utility of population-wide electronic health databases for risk stratification across cancer types has not been fully explored.

*Methods:* We use time-dependent Bayesian Cox Hazard models built on modern machine learning frameworks to scale the statistical approach to 6.7 million Danish individuals covering 193 million life-years over a period from 1978-2015. A set of 1,392 covariates from available clinical disease trajectories, text-mined basic health factors and family histories are used to train predictive models of 20 major cancer types. The models are validated on cancer incidence between 2015-2018 across Denmark and on 0.35 million individuals in the UK Biobank.

*Findings:* The predictive performance of models was found to exceed age-sex-based predictions in all but one cancer type. Models trained on Danish data perform similarly on the UK Biobank in a direct transfer without any additional retraining. Cancer risks are associated, in addition to heritable components, with a broad range of preceding diagnoses and health factors. The best overall performance was seen for cancers of the digestive system but also Thyroid, Kidney and Uterine Cancers. Risk-adapted cohorts may on average include 25% individuals younger than age-sex-based cohorts with similar incidence.

*Interpretation:* Data available in national electronic health databases can be used to approximate cancer risk factors and enable risk predictions in most cancer types. Model predictions generalise between the Danish and UK health care systems and may help to enable cancer screening in younger age groups.

*Funding:* Novo Nordisk Foundation.

**Research in Context:** *Evidence before this study:* A number of cancer risk prediction algorithms based on genetics or family history, lifestyle and health factors, as well as diagnostic tests have been developed to improve cancer screening by targeting individuals at increased risk. Many countries are assembling population-wide registries of electronic health records. Yet these resources do not necessarily encompass all the information required for currently available cancer risk models. It is therefore not clear yet how well national health data resources serve the purpose of population wide cancer risk prediction and cancer screening, which factors and data types are most informative for cancer specific and multi-cancer risk prediction and whether such algorithms would transfer between national health care systems.

*Added value of this study:* We developed risk prediction models for 20 major cancer types based on hospital admission records, family history of cancer cases, and some text-mined basic health factors across the Danish population from 1978 to 2015. The analysis shows that established and novel risk factors of different cancer types can be extracted from the vast amounts of data available in national health registries, facilitating accurate risk predictions. Further, validating the model on all adults residing in Denmark from 2015 to 2018 provides a unique opportunity to examine the potential of national-scale medical records for cancer risk prediction. Additionally, we validate the models in the UK Biobank, showing the transferability of the models across different health care systems. Lastly, we calculate that the information may facilitate earlier screening of individuals compared to an age-sex-based approach.

*Implications of all the available evidence:* Our study shows that national electronic health databases can help to identify individuals of increased risk of cancer across many organ sites. Model parameters approximate important cancer risk factors related to alcohol, smoking, metabolic syndromes and the female reproductive system. The ability to identify subsets of the population earlier compared to age-sex-based screening may improve the efficiency of current screening programs. The ability to predict a broad range of cancers may also benefit the implementation of new multi-cancer early detection tests, which are currently being trialled across the world.

## Introduction

Detecting cancer early can have a profound impact on treatment options and long-term survival rates across all cancer types ^1^. However, ∼50% of cancers are still diagnosed at a late stage ^2^. While early detection efforts, particularly national screening programs, have shown measurable improvements in health outcomes, they are currently only viable for a handful of organ sites, like Colorectal, Breast, Cervix Uteri, and Lung cancer ^3–6^.

The development of liquid biopsy tests for multi-cancer early detection ^7–10^ creates new possibilities for universal screening. While early clinical trial results show promising outcomes ^11^ and large-scale trials are currently conducted ^12^, risk adjusted targeting of the most susceptible individuals could further increase efficiency and enable broader implementation across age groups. Many bespoke risk models have been developed for specific cancer types including Colorectal ^13–15^, Melanoma ^16–18^, Lung ^19–21^ Prostate ^22–24^, Breast ^25–28^, Pancreatic ^29,30^, Liver ^31–33^, Stomach ^34,35^, Kidney cancer ^36^ or AML ^37^. The emergence of multi-cancer early detection tests warrant the development of pan-cancer risk models, which have recently been developed building on polygenic risk scores ^38^ or primary care data ^39^.

Cancer risk derives from many factors, ranging from environmental exposures ^40^, lifestyle choices ^41,42^ to inherited genetic predispositions ^43,44^. Even though these are not consistently available at an individual level across populations yet, many of the underlying factors are correlated and exhibit pleiotropic effects causing multiple cancer types ^45^, and other ailments ^46,47^. This provides an opportunity to approximate risk through data widely available in national health databases. With comprehensive data on a national scale becoming available in countries such as Denmark ^48–50^, or the United Kingdom ^51–53^ such opportunities become possible. However, the quantification of risk at a national level has not been comprehensively assessed and the transferability across different health care systems remains an open question.

Here, we make use of the Danish health registries to quantify the risks of 20 different cancer types based on prior diseases from secondary care, family history, and basic health factors for the majority of the population over 40 years. We validate these estimates in the UK Biobank to assess transferability across health care systems. Based on these risk estimates we assess the potential of population health data based cancer screening.

## Methods

### Data Sources

For this study, we make retrospective use of the Danish Health Registries, including the Central Person Registry (CPR), the Danish National Patient Registry (DNPR), the Death Registry (DR), the Cancer Registry (CR) and full text medical records from secondary care records in the BigTempHealth project ^54^, as the main data sources for model development and initial evaluation. Denmark constitutes a unique opportunity to study comorbidities and health-related factors with up to 40 years of linkable data collected in various registries across the entire population. Permissions for the work were obtained from the Danish Patient Safety Authority (3-3013-1731/1) and the Danish Health Data Authority (FSEID-00003092, FSEID-00003724, FSEID-00005633). A more detailed description of the used registries can be found in the supplementary materials.

An external validation cohort is provided through the UK Biobank, a cohort-based prospective study with roughly 500,000 participants aged 40-69 years when recruited between 2006 and 2010 in the United Kingdom ^55^. Access to the UK Biobank was granted under application 45761.

The study follows along with the transparent reporting of a multivariable prediction model for individual prognosis or diagnosis (TRIPOD) statement for reporting. The completed checklist can be found in the supplementary materials.

### Cohort

For model development on the Danish registries, we include all adults (16-86 years) without prior malignant cancers in the time period from January 01, 1995, to December 31, 2014. The dataset for development is randomly split into train (65%), validate (10%), and test (25%) sets. Prior malignancies are identified through any indication of a cancer case in any of the used registries. Additionally, individuals with information on only secondary/metastatic cancers or treatment for malignant cancers without information on the primary cancer are removed. Otherwise, no individuals are removed from consideration.

For model validation on the Danish registries, we consider all adults without a prior indication of cancer aged 16-75 years on January 01, 2015, and evaluate the model on cancer incidence from January 01, 2015, to April 10, 2018.

The external validation in the UK Biobank is similarly performed, except for an age range of 50-75 years, as the UK Biobank does not cover the full age spectrum.

### Features

In total we use 1,392 features of which 1,305 are binary, 84 are categorical, and 3 are continuous. All features contained in the model are allowed to vary over time, effectively incorporating the information in the registers as they become available. We use the last observation carried forward to model the progression in time.

For every eligible individual, we extract information on primary and secondary diagnoses from the DNPR. Diagnoses are coded as binary indicators, encoding if an individual ever had a certain disease at a given point in time. In total we have 1,305 disease indicators, corresponding to ICD-10 3rd level codes from chapters 1-18 (excluding C* - Malignant neoplasms).

Family history information is computed for each individual based on family trees of up to 2nd-degree relatives (except children of children). In total, we have 80 categorical variables encoding basic indicators for cancer cases in an individuals’ family for each of the 20 cancer types considered.

Basic health factors are extracted from doctor notes and cover aspects regarding alcohol consumption, smoking, height, weight, blood pressure, and age at first birth. Missing data are handled through dedicated indicators encoding the absence of information or via single mean imputations. An overview of the variables with a more detailed description of the used encodings is given in the supplementary table 1.

### Outcomes

The main outcome of interest for this study is the occurrence of a primary malignant cancer diagnosis. We focus on 20 major organ sites along with the groupings in NORDCAN ^56^. Additionally, to control for competing events, we analyse a composite outcome including all other malignant cancers (C*), and a non-cancer death outcome.

The date used for declaring a cancer diagnosis corresponds to the earliest indication of a malignant cancer in either DNPR, CR or DR. If there is a report of an unknown or uncertain cancer diagnosis (D37-49) and a malignant cancer is reported within the following year, the date of the uncertain diagnosis is used as the earliest indication. Additional details can be found in the supplementary materials.

### Model

The models underlying our analyses are based on a counting process representation of Cox’s partial-likelihood ^57,58^. We fit sex-stratified models to allow for different baseline hazards between the sexes. The time-axis for our models is age ^59^. Individuals come under risk when they reach the inclusion age or at the age at which they otherwise enter the population. Individuals are followed until the first occurence of cancer, death, emigration or the end of follow-up. The covariate effects are modelled through a linear predictor. To fit these models to big data, we use a Bayesian version of the Cox model as described in ^60^. The baseline hazard is estimated via Breslow’s estimator ^61^. Model predictions are based on the cumulative incidence function to account for competing events. We fit 22 cause-specific Cox models. Data from the year preceding an event or prior to the study end are removed to avoid identifying factors that are part of the diagnostic process. Hence, evaluations are generally based on predictions at least 1 year ahead. For details see the supplementary materials.

### Validation and evaluation

We evaluated our predictions on a test set covering 25% of the Danish population in the period from January 01, 1995, to December 31, 2014, in a similar setup as the initial development, e.g. dynamic risk assessment for 1-year ahead predictions. Additionally, we performed single time point evaluations to assess medium-time range predictions but also to further guarantee no possible time leakage.

We compute the risk for every eligible individual with covariate information up to January 01, 2014. Subsequently, these predictions are evaluated on cancer incidence from January 01, 2015, to April 10, 2018. To assess the discriminative power of our models we compute Harrell’s concordance index ^62^ and ROC curves. For the concordance index, we compute two different versions, one that incorporates age and sex explicitly and another where we evaluate the concordance index for sex-stratified data with age as the timeline, effectively comparing only individuals of the same sex across the same age, providing us with an indication of model performance conditional on age and sex.

To test for model improvements, we performed Likelihood Ratio tests adjusted for the Family Wise Error Rate (FWER) ^63^.

Further, we evaluate Kaplan-Meier (KM) curves for individuals in the top 1 risk percentile. The KM curve is then compared to a corresponding age-sex-based stratification. For the evaluation in Denmark, the baseline hazard is used as the age-sex comparator. For the UK Biobank evaluation, we compare our predictions to the baseline hazard estimate from Denmark but also on an additional age-sex Cox model (age up to a quadratic term + interactions) fitted on the UK Biobank data itself for a fairer comparison. Differences between the KM curves are assessed through a simple Cox regression (Hazard Ratio) and Log-Rank tests.

Calibration of our predictions is assessed for each decentile of the risk score. The observed rate is computed based on Kaplan-Meier estimates. Relative risk estimates are based on a age-sex matched cohort, where we select for each cancer case up to 100 healthy individuals with the same sex and +/-1 year of age.

To assess the screening performance we evaluate whether a risk-based redistribution of screening slots from an age-sex-based cohort identifies the same (or more) and also earlier cancer cases based on a similar number of individuals.

The analysis is based on 5 year age brackets with the starting age ranging from 40-65 in the Danish data and 55-65 in the UK Biobank in the validation period from 2015-2018. We evaluate the number of cancer cases within this cohort and their respective age distribution. Subsequently, we construct a similarly sized cohort, with the same female/male ratio based on the model risk scores for each cancer respectively.

## Results

The training cohort consists of the majority of the Danish population from 1978 to 2015 with text-mined health factors, detailed ICD-10 encoded disease trajectories, and family histories of different cancers (figure 1, table 1). The training data comprises 6,732,553 individuals covering 60 million hospital visits, 90 million diagnoses, a total of 193 million life-years and 444,835 cases of 20 different adult cancers. This information has been assembled into 1,392 time-dependent covariates for each individual. For validation, we cover 4,248,491 individuals with 67,401 cancer cases in Denmark and 377,004 individuals with 11,486 cancer cases in the UK Biobank. Additional characteristics of the cohorts are given in table 1 with age-sex incidence curves in the supplementary figures C1-20a. Further details about the training performance can be seen in supplementary figure 1.

**figure 1:**
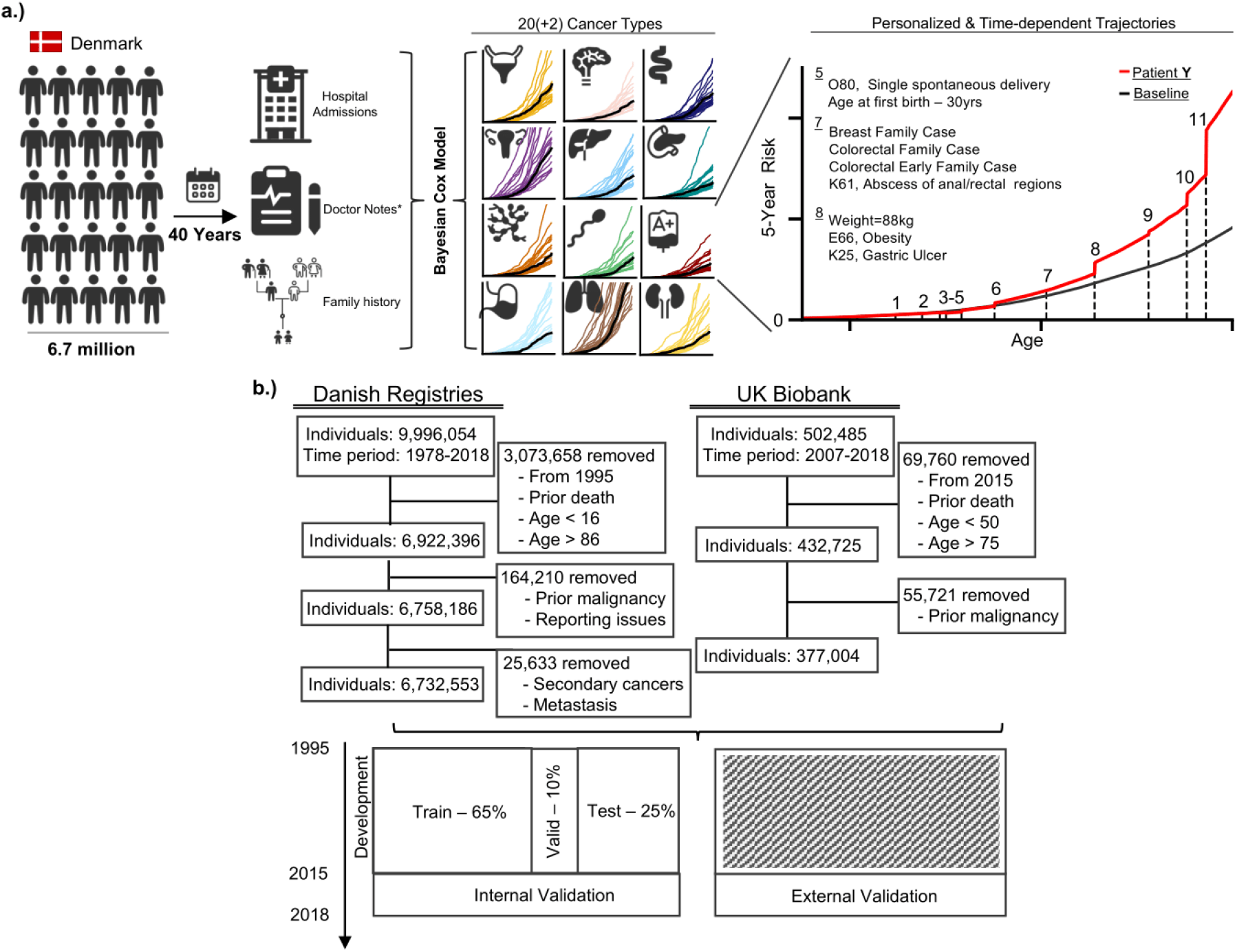
Study schematic and cohort overview. a) Schematic of the study. Electronic health records for 6.7 million Danes over 40 years are collected and used to build Bayesian Cox regression models for 20 cancer types plus 2 additional composite measures. These models can then be used to obtain dynamically evolving risk trajectories as exemplified here for a female patient Y. (1) N92, Exxcessive and irregular menstruation; (2) L20, Atopic dermatitis, Stomach Family Case; (3) O21, Excessive vomiting in pregnancy, O24, Diabetes mellitus in pregnancy; (4) N81, Female genital prolapse, J42, Unspecified chronic bronchitis; (5) O80, Single spontaneous delivery, Age at first birth – 30yrs; (6) Alcoholic, Non-Smoker, High Blood Pressure, No Low Blood Pressure; (7) Breast Family Case, Colorectal Family Case, Colorectal Early Family Case, K61, Abscess of anal/rectal regions; (8) Weight=88kg, E66, Obesity, K25, Gastric Ulcer; (9) E11, Type 2 diabetes mellitus, I70, Atherosclerosis, K20, Oesophagitis; (10) Breast Family case 1st degree, Breast Multiple Family Cases; (11) N61, Inflammatory disorders of breast b) Flow diagram of the sample selection process for the internal (Denmark) and external (UK Biobank) cohort. The bottom figure depicts the sample splits over the respective time periods

**table 1:**
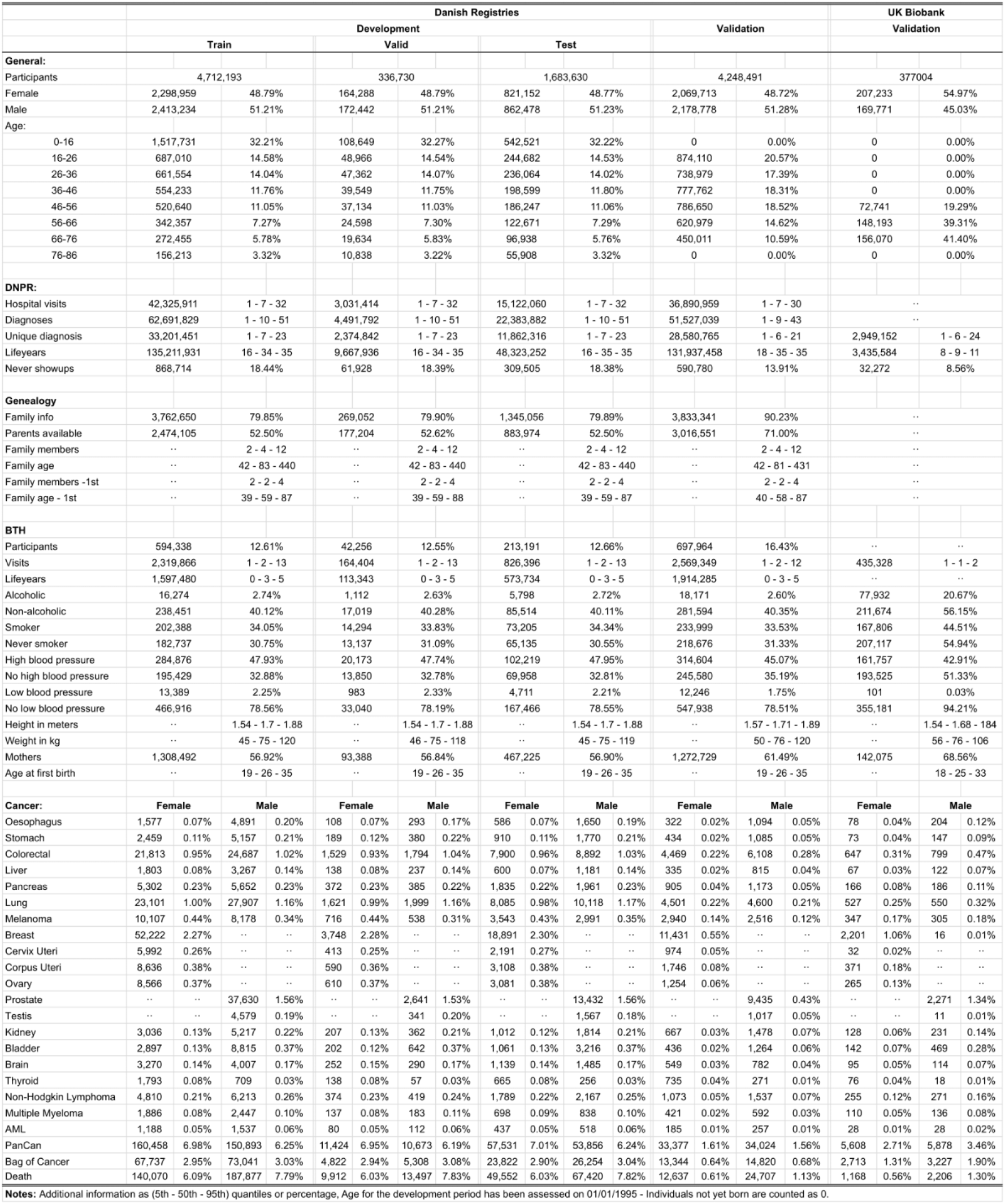
Cohort overview.

The national validation on the Danish population produced an average concordance index of 0·81 across cancer types, ranging from 0 ·66 (s.d.=0·007) for Cervix Uterine to 0·91 (s.d.=0·004) for Liver cancer (figure 2a). In total, 11 cancer sites have a concordance above 0·80 and 18 above 0·70 (supplementary table 2). There was a gained predictive value over standard age-sex based evaluations for all cancer sites except Ovary (log-rank test, FWER < 0·1, supplementary table 3) with age-sex adjusted concordance of on average 0·59 (range: 0·54-0·74, figure 2a). The top 1% risk quantile in each cancer constituted an average hazard ratio of 1·94 (range: 0·98 - 5·06) over age and sex, which was a significant improvement in 14 cancers with Thyroid (5·06, HPD90%: 2·97 - 8·65), Liver (4·12, HPD90%: 3·28 - 5·16), and Corpus Uteri (2·62, HPD90%: 1·94 - 3·52)) showing the largest risk spread (log-rank test, FWER < 0·1, supplementary figures C1-20e, supplementary table 4). Importantly, predicted and realised risks were well calibrated as measured across each decentile, demonstrating that the model produces meaningful estimates of the absolute cancer risks for each individual (figure 2b, supplementary figures C1-20d). For 10 cancer types with available QCancer risk models, the corresponding AUCs appear similar (supplementary table 5).

**figure 2:**
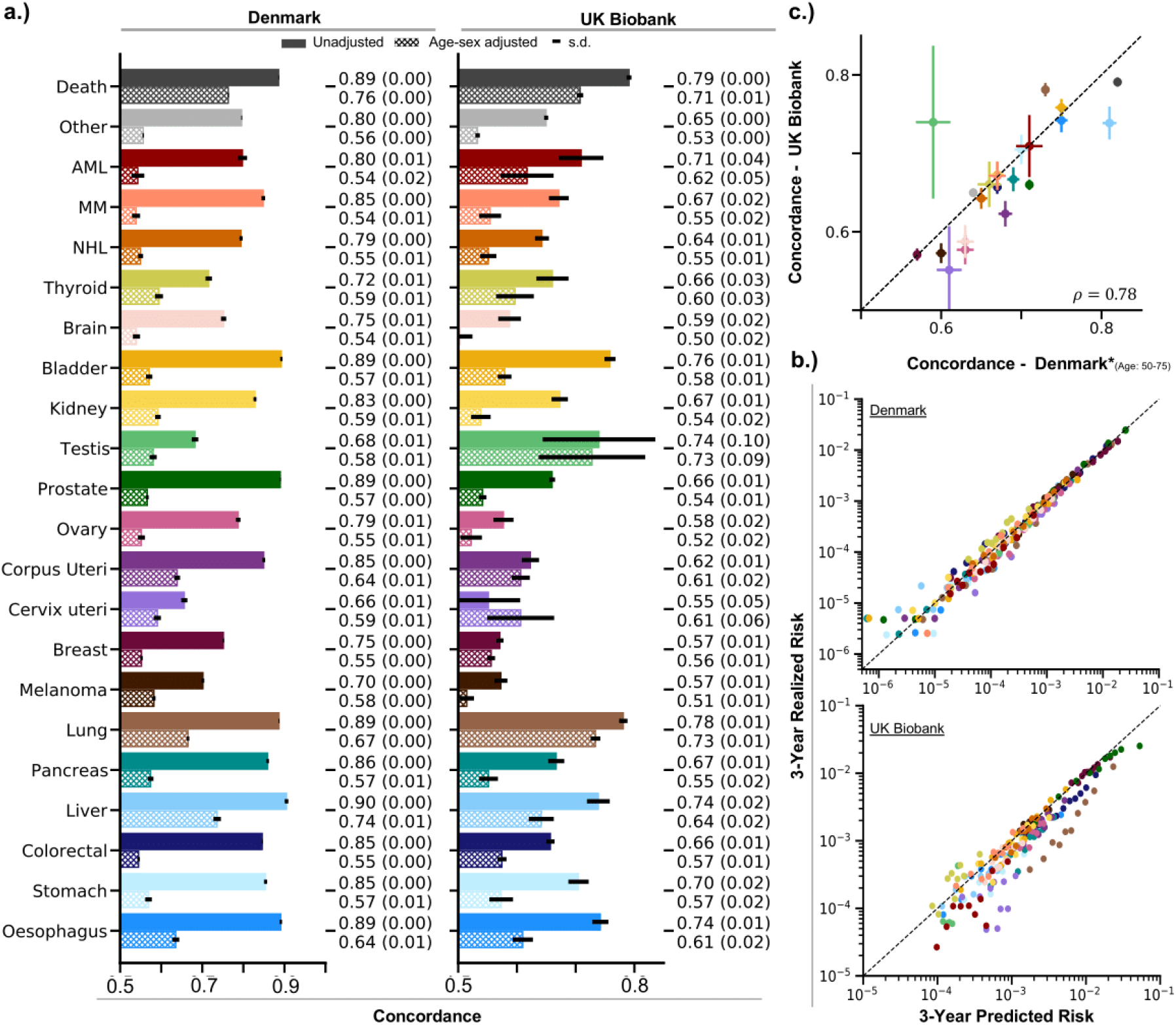
Risk prediction performance validation. a) Concordance index for each of the 22 possible outcomes evaluated from 2015 to 2018 based on risk predictions from 2014 across all of Denmark and the UK Biobank, respectively. The full bars correspond to a full model including age and sex while the hatched bar corresponds to the adjusted version, where we compare concordance within the same sex and across individuals of the same age. Standard deviations for the Concordance index are given in brackets. b) Calibration plots for the Danish and UK Biobank cohort, respectively. Calibration is evaluated for each risk decentile and compared to the corresponding observed rate based on KM estimates. c) Comparison of the Concordance index between the Danish estimates based on a similar age range (50-75) and the UK Biobank. Correlation is assessed via Pearsons’ ρ. Lines represent the standard deviation.

Cancer incidence in the UK Biobank from 2015 to 2018 was used to externally validate model predictions and to examine the international transferability between different health care systems. The average concordance index in the UK Biobank was 0·66, ranging from 0·55 (s.d.=0·054) for Cervix Uterine to 0·78 (s.d.=0·007) for Lung cancer. These values are comparable to the values for a corresponding age bracket of the Danish population (figure 2c). In total, 7 cancers have a concordance above 0·70 (figure 2a, supplementary table 2). Significant improvements over age and sex were found in 13 cancers (LR-Test, FWER < 0·1, supplementary table 3) with age-sex adjusted concordance of on average 0·59 (range: 0·50-0·73, figure 2a). The top 1% risk quantiles correspond to an average hazard ratio of 2·45 (range: 0·92 - 6·90), similar to the observations in Danish data and significant in 4 cancers (Liver, Lung, Breast, and Corpus Uteri; log-rank test, FWER < 0·1, supplementary table 4, supplementary figures C1-20e). The smaller number of significant effects is likely due to the smaller cohort size.

Calibration curves reveal a slight overestimation of the actual risk in the UK Biobank (figure 2b), which may be due to the healthy subject bias in the UK Biobank with roughly 12-18% lower cancer incidence than the general population ^64^. Overall these analyses show that health registry information can be used in risk models to quantify cancer risks for all major cancer types with competitive accuracy and transferability between Denmark and the UK.

Insights into the nature of risk predictions can be gained from the model’s hazard ratios. This enables us to explore whether the variables found to be associated with cancer risks correspond to known causal factors, are surrogates of other underlying risk factors or reflect the diagnostic pathway of suspected cancers.

Basic health factors contribute to almost all cancers with the highest number of associations found for Oesophagus, Breast and Melanoma. The most widely associated factors identified by our model are high alcohol consumption, Age at first birth and Height (figure 3a). Furthermore, we find a wide spectrum of diseases associated with the cancers studied. The cancer types with most associated diseases are Lung and Prostate while Multiple Myeloma and Testis show the fewest, based on hazard effects of at least 10% but also clear sign effects as evaluated by the highest posterior density 90% HPD. On average there are 28 associations (range: 6-122, figure 3a) with hazard effects of at least 10% and 13 with a clear sign effect (range: 3-75, figure 3a). The risk factors attributed by our models based on the 90% HPD for each cancer can be found in the forest plots in the supplementary figures C1-20c. The ICD-10 chapters with most associations over all cancers span the digestive, genitourinary, circulatory and musculoskeletal systems as well as precursor neoplasias. Family history is a contributing factor for all cancers with the most associations found in Colorectal, Melanoma and Testis. For 10 of the studied cancers we also see clear sign effects, covering mostly cancers with known heritability. Most widely associated are family cases of Lung, Breast, Prostate and Colorectal cancer, which are also found to be associated with elevated risks of other cancer types.

**figure 3:**
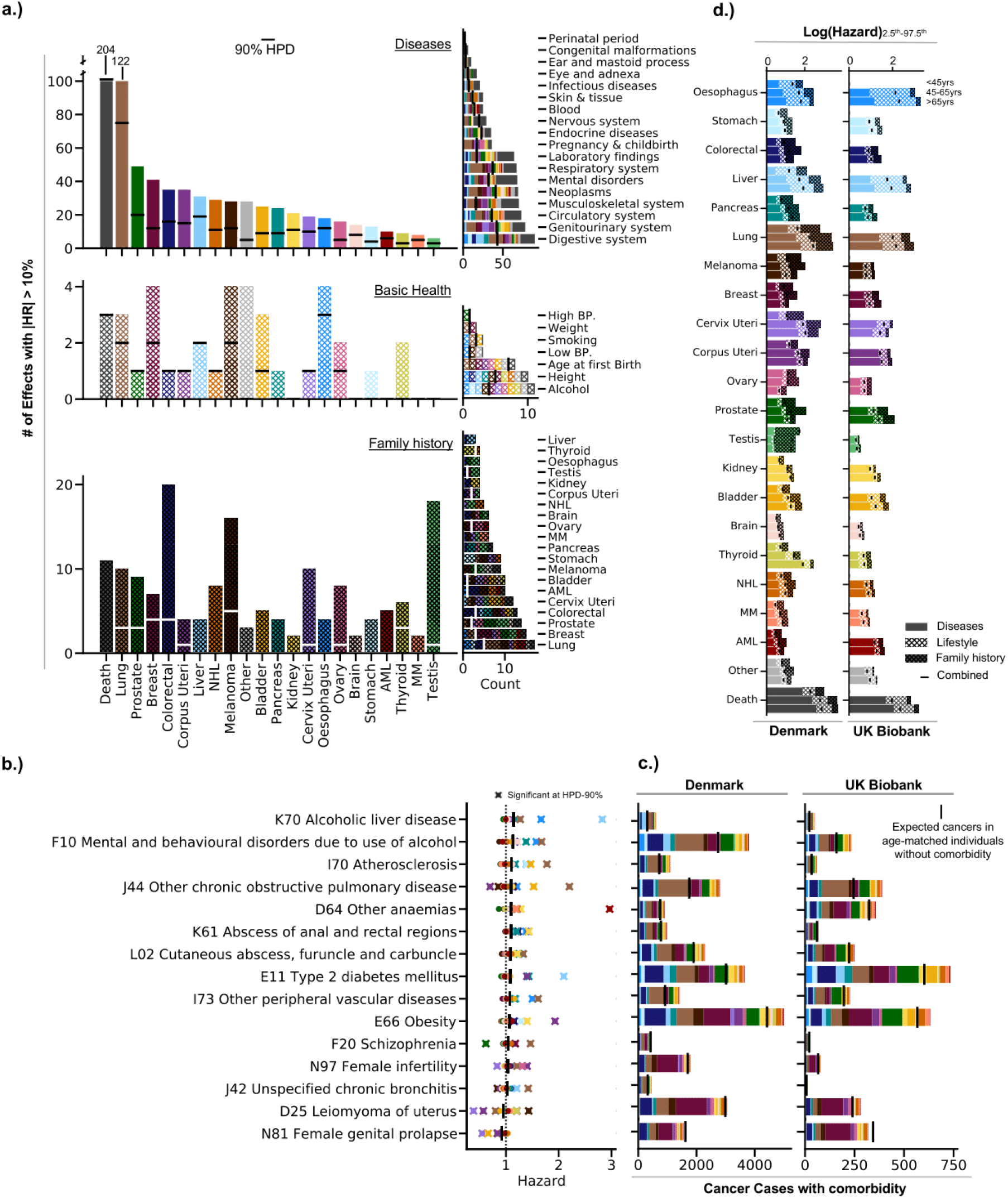
Attribution of risk factors. a) Count of associations with a hazard effect of at least 10% for each cancer type split by the 3 main covariate types Diseases, Basic health and Family history. The black/white bars correspond to an association that shows a clear sign effect based on the 90% HPD interval. The graphics to the right aggregate the association info by the covariate type rather then the individual cancers (ex. diseases in an ICD-10 chapter, # associations for Alcohol, # associations for family cases of Lung cancer). b) Covariates with at least 3 associations based on the 90% HPD. c) Number of cancer cases with the presence of a particular disease for Denmark and the UK Biobank, respectively. The black bar indicates the expected number of cancer cases based on relative risk estimates if the disease would appear at the same frequency in individuals with cancer as it does in otherwise healthy individuals. A black bar lower than the number of cases indicates a disease with a higher frequency in individuals with cancer while a higher bar indicates a lower frequency. d) Spread of the Log-Hazard between the 2.5th and the 97.5th percentile by the 3 main covariate types, evaluated for 3 different age brackets (<45 yrs, 45-65 yrs, and >65 yrs) for the Danish and UK Biobank cohort, respectively.

Several ICD-10 diagnoses are associated with multiple cancer types (figure 3b, HPD >=90% for 3 or more cancer types). The found diagnoses reflect canonical cancer risk factors, enabling approximate estimation thereof. Some of the emerging patterns include alcohol consumption represented by alcoholic liver disease (K70) or alcohol related mental disorders (F10) but also smoking through COPD (J44) and chronic bronchitis (J42). Interestingly, we can also identify metabolic syndromes like Obesity (E66) and Diabetes (E11) reflecting more diffuse lifestyle patterns of activity and diet, consequently leading to diseases like atherosclerosis (I70) and peripheral vascular disease (I73). We also identify various other factors like abscesses (K61, L02), anaemias (D64) but also schizophrenia (F20). Another important group are female reproductive diseases (N97, D25, N81) reflecting hormonal aspects in cancer. A full table of estimates for all 1,305 ICD-10 codes for each of the cancers considered can be found in the online materials.

To further assess the estimated effect sizes, we compute relative risk estimates for each diagnosis between cancer cases and age-sex matched controls. Reassuringly, we observe an enrichment or depletion for the aforementioned diagnoses (figure 3c). Comparable trends are observed in the UK Biobank, demonstrating that the found associations, irrespective of the underlying mechanism, tend to transfer between the two health care systems.

The overall contribution to each health data category on the risk within the population can be measured by the spread of the log-hazard derived from the variables and their effects. In both Danish and UK Biobank data disease histories have the strongest contribution with an increasing trend for older individuals – potentially because more diagnoses accrue over a lifetime, but possibly also reflecting differences in the aetiology of early and late onset disease (figure 3d). The contribution of basic health factors and family history are more variable and depend on the individual cancers.

One of the main aims of cancer risk prediction is improving early detection and cancer screening. As the incidence of many cancers rises as a power of age, screening is typically offered to individuals older than a certain age threshold and those with known predisposition.

Here we explore the potential of screening based on population health registries to assemble a risk informed cohort and compare it to a standard age-sex-based thresholding approach for 16 cancers, respectively. We have removed cancers of the Testis, Cervix uterine, and Thyroid along with Melanoma as the general population incidence shows decreasing or stagnating trends at certain age brackets. We evaluate whether a risk-based redistribution of screening slots from an age-sex-based cohort identifies the same (or more) and also earlier cancer cases based on a similar number of individuals.

Results for all age brackets can be found in the supplementary table 6. For the age bracket from 55-60, which could be evaluated in both cohorts, the risk based screening contains a similar number of cancer cases. There are on average 12% more cancer cases for the Danish cohort (range: -1% to 37%, figure 4) with all except Ovary and Non-Hodgkin Lymphoma showing an improvement. The best performing cancer sites are Liver (1·37, s.d=0·10) and Corpus Uteri (1·22, s.d.=0·08). In the UK Biobank the risk based cohort contains on average 7% more cases (range: -14% to 32%, figure 4) with 12 cancer sites showing an improvement and Lung (1·32, s.d.=0·15) along with Corpus Uteri (1·16, s.d.=0·18) as the best performing cancer sites.

**figure 4:**
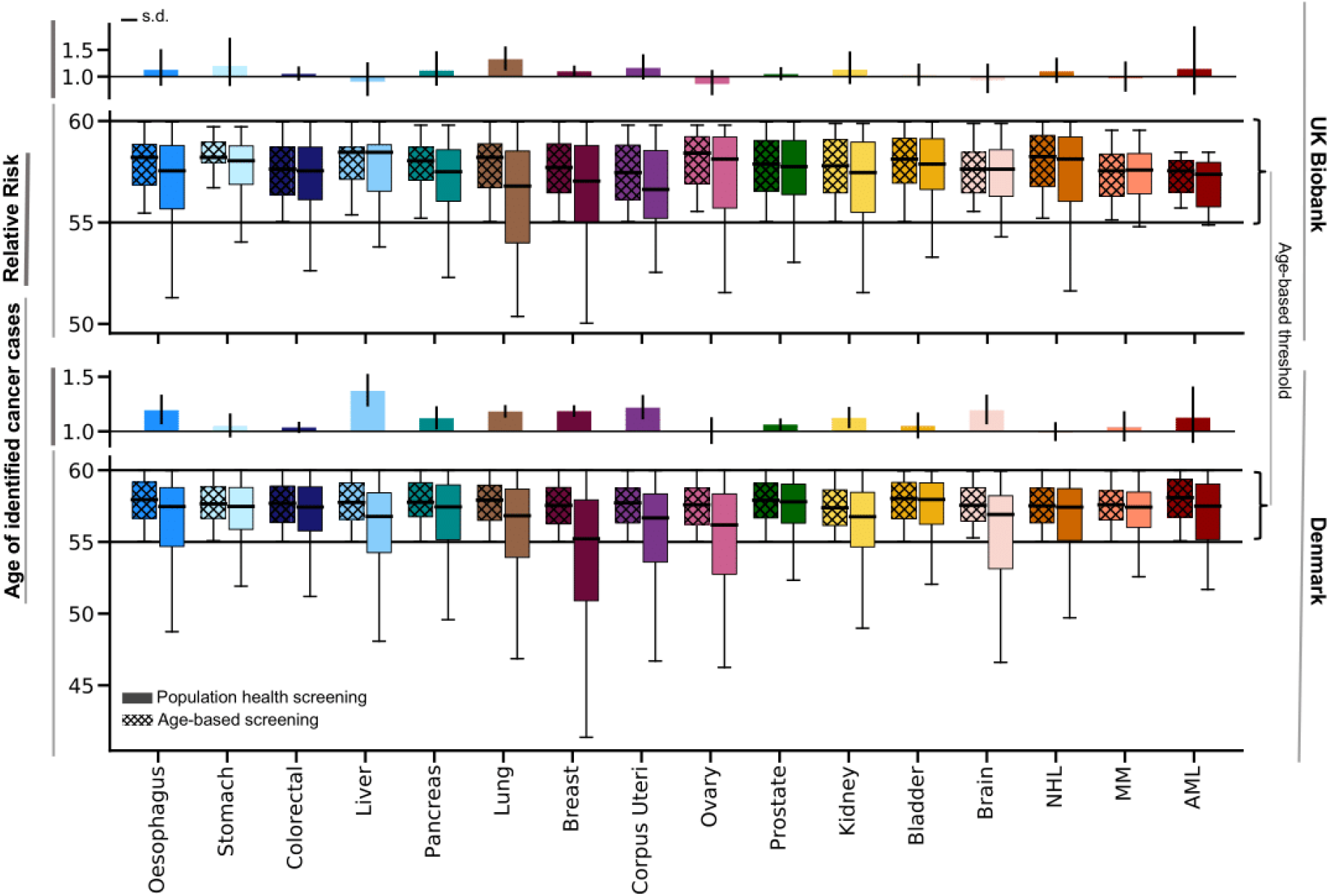
Screening evaluation Age distribution of screening cohorts compared between an age-sex-based cohort (55-60 years of age) versus a similarly structured risk based cohort for each of the 16 cancers, respectively. Relative risk estimates are computed for the number of identified cancers between the respective screening regimes.

The age distribution of the cancer cases in the risk based cohort shows a clear shift towards younger individuals for most cancer sites.

The mean age of individuals with cancer is on average 1·49 years younger in Denmark (range: 0·34-3·7, figure 4) and 0·67 years in the UK Biobank (range: 0·03-1·58, figure 4). Further examining the age distribution reveals that in Denmark on average 26% (range: 12% to 48%, figure 4) of the respective cancer cases are younger than the corresponding age threshold, with 11 cancer sites above 20%.

In the UK Biobank, on average 17% of the identified cancer cases are younger than the age threshold (range: 4% to 33%, figure 4) with 6 cancer sites above 20%. Oesophagus, Lung, Breast, Corpus Uteri, and Non-Hodgkin Lymphoma have 20% of the identified cancer cases younger than the age threshold in both cohorts, highlighting the overall shift in the age distribution.

Other age brackets provide similar results (supplementary table 6).

While the translation into targeted screening programmes would need to be assessed by randomised trials, our results show that there is the potential to utilise cancer risk models to assemble younger cohorts with the same or higher incidences. It is plausible yet unproven that these benefit from earlier intervention.

## Discussion

The analysis showed that cancer risk predictions on population health data resources are possible across most cancer types, are transferable between Danish and UK health care systems and likely to enable screening younger population cohorts with a similar number of incident cancers.

The factors identified by the models are not necessarily causal and should thus be interpreted with care. While many diagnoses were found to be associated with differential cancer risks these findings may just reflect other underlying behavioural risk factors, predispositions, or the diagnostic pathway of cancer detection.

Another limitation pertains to the assessment of cancer screening. The ability to analyse younger cohorts does not imply an ability for earlier detection, as this depends on the characteristics of the detection assay. Rather, our analysis shows that it may be possible to redistribute tests to increase the efficiency of screening. As a next step it may be instructive to conduct a retrospective analysis of currently ongoing multi-cancer early detection trials to assess whether detactable positive cases are exhibiting elevated risks as quantified here. The Danish population health data resources are unique as they cover a considerable period and can be linked to each other. Yet basic health parameters were derived from text mining of medical records available for only a subset of the population. Also, the data derived from the hospital system by definition only cover those to happen to be in contact with a hospital. This limits data availability and quality and in many instances misses important information on risk factors such as smoking or alcohol consumption. The systematic missingness of data may also increase the risk of algorithmic bias and any personalised screening implementation should thus evaluate fairness.

With efforts to build national digital health infrastructures a question arises which type of data would be best suited for the purpose of cancer risk assessment. The analysis showed that ICD-10 diagnostic codes from secondary care are well suited and transfer across these two European health care systems and can efficiently be used to approximate important risk factors. Yet basic health data text mined from secondary care records also contributed important information in line with previous investigations. It therefore appears beneficial to gather such information for a broad set of the population ideally with further emphasis on known behavioural risk factors, notwithstanding the difficulties of their quantification. Lastly, family history of diverse cancer types was a measurable risk factor for a range of cancers. While these data could be derived due to the specifics of the Danish disease and civil registration registries, data protection issues may make it harder in other countries. An alternative and further opportunities may arise from including genome sequencing data. Lastly, it is important to recognise that the information used is equitable and does not exacerbate existing health disparities.

Taken together, our analysis shows that cancer risk prediction based on population health data resources is possible and suggests a benefit for and road towards risk adapted cancer screening.

## Data Availability

Danish registry data are available for use in secure, dedicated environments via
application to the Danish Patient Safety Authority and the Danish Health
Data Authority.
UK Biobank data are available to verified researchers on application at http://www.UK Biobankiobank.ac.uk/using-the-resource/.
Code is available on https://github.com/gerstung-lab/CancerRisk

https://github.com/gerstung-lab/CancerRisk

## Contributors

AWJ developed the risk prediction model, conducted Danish and UK Biobank data analyses, assembled all figures and wrote the manuscript with MG. PCH analysed Danish secondary care records. KG helped with UK Biobank data analysis. JXH and DP helped with data processing. LHM advised on statistical and epidemiological aspects. EB and SB coordinated data collection. MG conceived the study and supervised the analysis. AWJ and SB accessed and verified the Danish data and AWJ and MG did so for the UKB. All authors approved the manuscript.

## Acknowledgements

This work was supported by grant NNF17OC0027594 from the Novo Nordisk Foundation.

## Data sharing

Danish registry data are available for use in secure, dedicated environments via application to the Danish Patient Safety Authority and the Danish Health Data Authority.

UK Biobank data are available to verified researchers on application at http://www.UKBiobankiobank.ac.uk/using-the-resource/.

Code is available on https://github.com/gerstung-lab/CancerRisk

