## Supplementary Materials for "Multi-cancer risk stratification based on national health data: A retrospective modelling and validation study"

### Table of Contents

**Supplementary Materials** **1**
Additional Methods 2
Danish Registries 2
Outcomes 2
Model Description 2
Validation and Evaluation 3
Additional Figures and Tables 6
ST1: Features overview. 6
ST2: Additional metrics for the validation. 7
ST3: Model comparison. 8
ST4: Kaplan Meier curve comparison of top risk quantiles between our predictions and age-sex alone. 9
ST5: Performance comparison to QCancer 10
ST6: Screening evaluation 11
ST7: Baseline risk comparison to Age-sex model on the UK Biobank 13
ST8: Proportional hazards assumption for the overall predictor. 14
SF1: Age-sex adjusted concordance for the development data. 15
SFC-X: Summary plots 15
SFC1: Oesophagus 16
SFC2: Stomach 17
SFC3: Colorectal 18
SFC4: Liver 19
SFC5: Pancreas 20
SFC6: Lung 21
SFC7: Melanoma 22
SFC8: Breast 23
SFC9: Cervix Uteri 24
SFC10: Corpus Uteri 25
SFC11: Ovary 26
SFC12: Prostate 27
SFC13: Testis 28
SFC14: Kidney 29
SFC15: Bladder 30
SFC16: Brain 31
SFC17: Thyroid 32
SFC18: Non-hodgkin lymphoma 33
SFC19: Multiple Myeloma 34
SFC20: AML 35

### Supplementary Materials

#### Additional Methods

##### Danish Registries

All registers are linkable through a unique personal identification number provided in the Central Person Registry (CPR). We use the Danish National Patients Registry (DNPR) [65], a nationwide longitudinal register with data on hospital admissions (secondary care) across all of Denmark since 1977, to extract information on individual diagnoses.
The Danish Register of Causes of Death (DR) [66], covering all citizens dying in Denmark since 1970, and the Danish Cancer Register (CR) [67], collating cancer incidence across the Danish population from 1978 onwards, are used to supplement the information in the DNPR.
Additionally, we have been using the BigTempHealth project to extract information about basic health factors through doctor notes from hospitals in Region Zealand and the Capital Region since September, 2004.

##### Outcomes

As mentioned in the main text, we focus on 20 major organ sites along with the groupings in NORDCAN [56] including: Oesophagus (C15), Stomach (C16), Colorectal (C18-21), Liver (C22), Pancreas (C25), Lung (C33-34), Melanoma (C43), Kidney (C64), Bladder (C67), Brain (C71), Thyroid (C73), Non-Hodgkin Lymphoma (C82-86), Multiple Myeloma (C90), AML (C*- 920, 924, 925, 926, 928, 930, 940, 942) for both sexes. Female-only cancers include: Breast (C50), Cervix Uteri (C53), Corpus Uteri (C54), and Ovary (C56). Male-only cancers include: Prostate (C61), Testis (C62).
Cancers are identified by an entry in either the DNPR (primary + secondary diagnosis), CR, or DR. If multiple cancers are reported within a 3-month time window, all the corresponding cancers in the time window are considered equally valid as the primary.
For cancers reported in the CR, we check the morphological codes for benign, in situ, and metastatic cancers which are considered invalid.
The 20 major organ sites are preferentially treated to the bag-of-cancer and death outcome. If multiple cancers are considered, the earliest date is used for both.

##### Model Description

As mentioned we make use of the method developed in [60]. However, as there are 20 different cancer types of interest + 2 additional composite outcomes, we expand the approach to account for competing risks.
We fit 22 cause-specific Cox models on the development-train set of the Danish registries, censoring individuals when they transition to a competing state. The validate set is used to gauge some of the hyperparameters and for initial model evaluation and sensitivity assessment. The test set has not been part of any model derivation and is solely used for final model evaluations.
The cause specific hazard function $${\alpha_k(t) = \lim_{\Delta t \to 0} (\Delta t)^{-1} \mathcal{P}(T \leq t + \Delta t \hspace{0.05cm}, K=k | \hspace{0.05cm} T > t)}$$

for an event k can be written under the cox model as $$\alpha_k(t) = \alpha_{0k}(t) exp \big[ \boldsymbol{\theta}_k^{\top}\mathbf{X}(t) \big] $$, where $$\theta$$ are the model parameters and $$X(t)$$ are the time-varying covariates. The cause-specific cumulative hazard follows as $${A_k(t) = \int_0^t \alpha_k(u)\textrm{d}u}$$ and is estimated via Breslow's estimator [61] . The overall survival function from any of the possible competing causes follows as $$S(t) = exp\big[-\sum_{k=1}^{22} A_k(t) \big]$$. We can then define the cumulative incidence function (CIF) as $${F_k(t) = \int_0^t \alpha_k(u)S(u)\textrm{d}u}$$, incorporating the cause-specific hazards for each of the 22 event types [68]. The CIF forms the basis for most of your predictions. It should be noted that survival probabilities with internal time-varying covariates are ill-defined; we do assume here for simplicity that covariates stay constant over the prediction horizon.

For the final model specification, we used a StudentT(loc=0, scale=0.01) distribution with 1.5 degrees of freedom as prior and a lower rank(50) Multivariate-Normal distribution as the distributional family for stochastic variational inference. Overall, we use a batch size of 720 randomly selected individuals with a 1/20 ratio between events and non-events for each stochastic gradient descent update.

#### Validation and Evaluation

In total, we compared 6 different settings per cancer. The full model prediction is compared to an age-sex model based on the estimates from Denmark. A similar evaluation but now with an age-sex model (age up to a quadratic term and interactions) fitted on the UK Biobank data itself. We assessed the compatibility of the age-sex models and generally, the models perform similarly based on the ROC-AUC. Additionally, we evaluated Spearman's rank correlation between the two scores and almost all show a $$\roh > 0.9$$ except for Breast (0.62), Thyroid (0.74), and Melanoma (0.84) (supplementary table 7).

Testis and Cervix Uteri show a reversed correlation indicating a particular discordance between the underlying incidence curves.

Additionally, we evaluated the model improvements for incorporating the family history data and/or the health data (disease + basic health) individually. The combined predictor is assessed for the proportional hazards assumption (supplementary table 8).

Further, we stratify individuals based on their predicted risk of the top 1% quantile and assess the corresponding Kaplan-Meier (KM) curve. The KM curve is then compared to a corresponding age-sex-based stratification. We use the additionally fitted age-sex model for the UK Biobank. Differences between the KM curves are assessed through a simple Cox regression (Hazard Ratio) and Log-Rank tests (supplementary table 4, supplementary figure C1-20e). Calibration of our predictions is assessed for each decentile of the risk score.

TRIPOD Checklist: Prediction Model Development

| Section/Topic | Item | Checklist Item | Page |
|---|---|---|---|
| **Title and abstract** | | | |
| Title | 1 | Identify the study as developing and/or validating a multivariable prediction model, the target population, and the outcome to be predicted. | p.1 |
| Abstract | 2 | Provide a summary of objectives, study design, setting, participants, sample size, predictors, outcome, statistical analysis, results, and conclusions. | p. 2 |
| **Introduction** | | | |
| Background and objectives | 3a | Explain the medical context (including whether diagnostic or prognostic) and rationale for developing or validating the multivariable prediction model, including references to existing models. | p. 3-4 |
| | 3b | Specify the objectives, including whether the study describes the development or validation of the model or both. | p. 3 |
| **Methods** | | | |
| Source of data | 4a | Describe the study design or source of data (e.g., randomized trial, cohort, or registry data), separately for the development and validation data sets, if applicable. | p.4-5<br>24 |
| | 4b | Specify the key study dates, including start of accrual; end of accrual; and, if applicable, end of follow-up. | p. 4<br>fig. 1b |
| Participants | 5a | Specify key elements of the study setting (e.g., primary care, secondary care, general population) including number and location of centres. | p.4-5<br>24 |
| | 5b | Describe eligibility criteria for participants. | p. 5<br>fig. 1b |
| | 5c | Give details of treatments received, if relevant. | - |
| Outcome | 6a | Clearly define the outcome that is predicted by the prediction model, including how and when assessed. | p.6<br>p. 24 |
| | 6b | Report any actions to blind assessment of the outcome to be predicted. | p. 6 |
| Predictors | 7a | Clearly define all predictors used in developing or validating the multivariable prediction model, including how and when they were measured. | p.5-6<br>s. tab. 1 |
| | 7b | Report any actions to blind assessment of predictors for the outcome and other predictors. | p. 6 |
| Sample size | 8 | Explain how the study size was arrived at. | p.5<br>fig. 1b |
| Missing data | 9 | Describe how missing data were handled (e.g., complete-case analysis, single imputation, multiple imputation) with details of any imputation method. | p.5-6<br>s. tab. 1 |
| Statistical analysis methods | 10a | Describe how predictors were handled in the analyses. | p.5-6<br>p. 25<br>s. tab. 1 |
| | 10b | Specify type of model, all model-building procedures (including any predictor selection), and method for internal validation. | p.5-7<br>p. 25 |
| | 10d | Specify all measures used to assess model performance and, if relevant, to compare multiple models. | p.6-7<br>p.25 |
| Risk groups | 11 | Provide details on how risk groups were created, if done. | p.6-7<br>p.25 |
| **Results** | | | |
| Participants | 13a | Describe the flow of participants through the study, including the number of participants with and without the outcome and, if applicable, a summary of the follow-up time. A diagram may be helpful. | p. 5<br>fig. 1b |
| | 13b | Describe the characteristics of the participants (basic demographics, clinical features, available predictors), including the number of participants with missing data for predictors and outcome. | tab. 1 |
| Model development | 14a | Specify the number of participants and outcome events in each analysis. | tab. 1 |
| | 14b | If done, report the unadjusted association between each candidate predictor and outcome. | online m. |

| | | | |
|---|---|---|---|
| Model specification | 15a | Present the full prediction model to allow predictions for individuals (i.e., all regression coefficients, and model intercept or baseline survival at a given time point). | online m. |
| | 15b | Explain how to the use the prediction model. | online m. |
| Model performance | 16 | Report performance measures (with CIs) for the prediction model. | p.7-10 fig 1 |
| **Discussion** | | | |
| Limitations | 18 | Discuss any limitations of the study (such as nonrepresentative sample, few events per predictor, missing data). | p. 10-11 |
| Interpretation | 19b | Give an overall interpretation of the results, considering objectives, limitations, and results from similar studies, and other relevant evidence. | p. 10-11 |
| Implications | 20 | Discuss the potential clinical use of the model and implications for future research. | p. 10-11 |
| **Other information** | | | |
| Supplementary information | 21 | Provide information about the availability of supplementary resources, such as study protocol, Web calculator, and data sets. | p.12 |
| Funding | 22 | Give the source of funding and the role of the funders for the present study. | p.2 |

We recommend using the TRIPOD Checklist in conjunction with the TRIPOD Explanation and Elaboration document.

### Additional Figures and Tables

#### **ST1:** Features overview.

| Category | Variable | Count | Encoding | Missingness | Transformations | Data Sources | | Definition |
|---|---|---|---|---|---|---|---|---|
| | | | | | | Danish Registries | UK Biobank | |
| **Disease codes:** | | | | | | | | |
| | Certain conditions originating in the perinatal period | 59 | binary - (0, 1) | ·· | ·· | DNPR | Cat.: 1712 | A total of 1305 diseases corresponding to ICD-10 3rd level codes from chapters 1-18 (excluding C*) are extracted from the DNPR (primary and secondary diagnoses only) and coded as binary indicators (0, 1), representing whether an individual ever had the disease by a given point in time. Diagnoses before 1995 were encoded in ICD-8 and have been transformed to ICD-10 based on previously used mappings. We rely on category 1712 for the external validation in the UKB, which is a composite of hospital admission records, primary care records, and self-reported diseases. The chapters 'Neoplasm' and 'Symptoms, signs and abnormal clinical and laboratory findings, not elsewhere classified' are adapted from category 2000 (Hospital Episodes) as they are not provided in the other category. |
| | Certain infectious and parasitic diseases | 173 | binary - (0, 1) | ·· | ·· | DNPR | Cat.: 1712 | |
| | Congenital malformations, deformations and chromosomal abnormalities | 87 | binary - (0, 1) | ·· | ·· | DNPR | Cat.: 1712 | |
| | Diseases of the blood and blood-forming organs and certain disorders involving the immune mechanism | 34 | binary - (0, 1) | ·· | ·· | DNPR | Cat.: 1712 | |
| | Diseases of the circulatory system | 77 | binary - (0, 1) | ·· | ·· | DNPR | Cat.: 1712 | |
| | Diseases of the digestive system | 72 | binary - (0, 1) | ·· | ·· | DNPR | Cat.: 1712 | |
| | Diseases of the ear and mastoid process | 24 | binary - (0, 1) | ·· | ·· | DNPR | Cat.: 1712 | |
| | Diseases of the eye and adnexa | 47 | binary - (0, 1) | ·· | ·· | DNPR | Cat.: 1712 | |
| | Diseases of the genitourinary system | 82 | binary - (0, 1) | ·· | ·· | DNPR | Cat.: 1712 | |
| | Diseases of the musculoskeletal system and connective tissue | 79 | binary - (0, 1) | ·· | ·· | DNPR | Cat.: 1712 | |
| | Diseases of the nervous system | 68 | binary - (0, 1) | ·· | ·· | DNPR | Cat.: 1712 | |
| | Diseases of the respiratory system | 64 | binary - (0, 1) | ·· | ·· | DNPR | Cat.: 1712 | |
| | Diseases of the skin and subcutaneous tissue | 72 | binary - (0, 1) | ·· | ·· | DNPR | Cat.: 1712 | |
| | Endocrine, nutritional and metabolic diseases | 74 | binary - (0, 1) | ·· | ·· | DNPR | Cat.: 1712 | |
| | Mental and behavioural disorders | 78 | binary - (0, 1) | ·· | ·· | DNPR | Cat.: 1712 | |
| | Neoplasms | 48 | binary - (0, 1) | ·· | ·· | DNPR | Cat.: 2000 | |
| | Pregnancy, childbirth and the puerperium | 76 | binary - (0, 1) | ·· | ·· | DNPR | Cat.: 1712 | |
| | Symptoms, signs and abnormal clinical and laboratory findings, not elsewhere classified | 91 | binary - (0, 1) | ·· | ·· | DNPR | Cat.: 2000 | |
| **Genealogy:** | | | | | | | | |
| | Family case | 20 | categorical - (-1, 0, 1) | Effect coding: missing := 0 | ·· | CPR, DNPR, CR, DR | Cat.: 20107, 20110, 20111 | Genealogy information is computed for each individual based on information of up to 2nd-degree relatives (excluding the children of children). Families are reconstructed based on information on the father and mother from the CPR. Information on half-siblings is considered, while zygosity information for twins was not available. We use effect coding for the indicators and add a state indicating missingness. Effectively, -1 encodes no effect/not present, 0 encodes missingness (here: if there is no information on any of the parents), and 1 for effect/present.<br>For the UKB analysis, we rely on categories 20107, 20110, and 20111. However, genealogy information is only provided for first-degree relatives with Prostate, Breast, Bowel, and Lung cancers. For other cancers and the early cancer indicator, we set the covariates to 0, indicating insufficient information on family history. |
| | Family case (1st degree relatives) | 20 | categorical - (-1, 0, 1) | Effect coding: missing := 0 | ·· | CPR, DNPR, CR, DR | Cat.: 20107, 20110, 20111 | |
| | Early age (age <= 50 years) Family case | 20 | categorical - (-1, 0, 1) | Effect coding: missing := 0 | ·· | CPR, DNPR, CR, DR | ·· | |
| | Multiple family cases | 20 | categorical - (-1, 0, 1) | Effect coding: missing := 0 | ·· | CPR, DNPR, CR, DR | Cat.: 20107, 20110, 20111 | |
| **Basic Health:** | | | | | | | | |
| | Alcoholic | 1 | categorical - (-1, 0, 1) | Effect coding: missing := 0 | ·· | BTH, DNPR | Cat.: 1558 | Indicator of "ever had a problematic alcohol consumption", defined as either directly noted in the doctor's notes, as drinking 20 units per week or having had an alcohol-related disease (F10*, G621, G312, G721, I426, K70*, K292, K860, Z502, Z714, Z721). No indication of an alcoholic problem is coded as -1 if doctor notes are available, 0 otherwise. If evidence of an alcoholic problem is found the indicator is set to 1. For the UKB validation, we use the survey question about alcoholic consumption in Cat. 1558 with "daily" encoded as 1, "almost daily" encoded as 0, and the rest encoded as -1. |
| | Smoking | 1 | categorical - (-1, 0, 1) | Effect coding: missing := 0 | ·· | BTH, DNPR | Cat.: 20116 | Indicator of "ever smoker" as mentioned in the doctor's notes or through codes (F17*, Z508, Z716, Z720). Similar coding as for alcoholic. For the UKB validation, we use the survey question about smoking behavior in Cat. 20116 with current or former smoker encoded as 1. |
| | High blood pressure | 1 | categorical - (-1, 0, 1) | Effect coding: missing := 0 | ·· | BTH, DNPR | Cat.: 4079, 4080 | Indicator of "ever had high blood pressure" as diastolic >=90 and systolic >=140 or (I10-13, I15). Similar coding as for alcoholic. [Units in mm Hg] |
| | Low blood pressure | 1 | categorical - (-1, 0, 1) | Effect coding: missing := 0 | ·· | BTH, DNPR | Cat.: 4079, 4080 | Indicator of "ever had low blood pressure" as diastolic <=60 and systolic <=90 or (I95). Similar coding as for alcoholic. [Units in mm Hg] |
| | Height | 1 | continuous (cm) | Single mean imputation: (165f, 177m). | centered : 170 scaled:100 | BTH | Cat.: 50 | The height is extracted from the doctor's notes. Missing values are single imputed by the corresponding mean for the sex (165f, 177m). |
| | Weight | 1 | continuous (kg) | Single mean imputation: (69f, 82m). | centered : 75 scaled:100 | BTH | Cat.: 21002 | The weight is extracted from the doctor's notes. Missing values are single imputed by the corresponding mean for the sex (69f, 82m). |
| | Age at first birth | 1 | continuous (years) | ·· | centered : - scaled:100 | CPR, DNPR | Cat.: 2754 | Age at first birth as extracted from the CPR (ID appears as the mother for someone else) or a child birth-related diagnosis in the DNPR (O81-84, Z37, Z39) |

**ST2:** Additional metrics for the validation.

| | Cancer | Concordance | s.d. | AUC | Brier score |
|---|---|---|---|---|---|
| Denmark | Oesophagus | 0.891 | 0.003 | 0.889 | 0.000 |
| | Stomach | 0.854 | 0.004 | 0.852 | 0.000 |
| | Colorectal | 0.846 | 0.001 | 0.845 | 0.002 |
| | Liver | 0.905 | 0.004 | 0.903 | 0.000 |
| | Pancreas | 0.859 | 0.003 | 0.857 | 0.000 |
| | Lung | 0.887 | 0.001 | 0.885 | 0.002 |
| | Melanoma | 0.702 | 0.003 | 0.703 | 0.001 |
| | Breast | 0.751 | 0.002 | 0.753 | 0.005 |
| | Cervix Uteri | 0.656 | 0.007 | 0.659 | 0.000 |
| | Corpus Uteri | 0.850 | 0.003 | 0.849 | 0.001 |
| | Ovary | 0.787 | 0.005 | 0.786 | 0.001 |
| | Prostate | 0.890 | 0.001 | 0.890 | 0.004 |
| | Testis | 0.682 | 0.008 | 0.691 | 0.000 |
| | Kidney | 0.828 | 0.003 | 0.827 | 0.001 |
| | Bladder | 0.892 | 0.003 | 0.890 | 0.000 |
| | Brain | 0.752 | 0.006 | 0.751 | 0.000 |
| | Thyroid | 0.716 | 0.008 | 0.718 | 0.000 |
| | Non-Hodgkin Lymphoma | 0.794 | 0.004 | 0.793 | 0.001 |
| | Multiple Myeloma | 0.848 | 0.004 | 0.846 | 0.000 |
| | AML | 0.798 | 0.011 | 0.797 | 0.000 |
| | Other | 0.796 | 0.001 | 0.796 | 0.007 |
| | Death | 0.886 | 0.001 | 0.887 | 0.008 |
| UK Biobank | Oesophagus | 0.742 | 0.014 | 0.739 | 0.001 |
| | Stomach | 0.705 | 0.017 | 0.701 | 0.001 |
| | Colorectal | 0.657 | 0.007 | 0.654 | 0.004 |
| | Liver | 0.739 | 0.019 | 0.736 | 0.001 |
| | Pancreas | 0.667 | 0.014 | 0.663 | 0.001 |
| | Lung | 0.781 | 0.007 | 0.779 | 0.003 |
| | Melanoma | 0.573 | 0.011 | 0.570 | 0.002 |
| | Breast | 0.571 | 0.006 | 0.571 | 0.011 |
| | Cervix Uteri | 0.551 | 0.054 | 0.550 | 0.000 |
| | Corpus Uteri | 0.623 | 0.015 | 0.622 | 0.002 |
| | Ovary | 0.577 | 0.017 | 0.576 | 0.001 |
| | Prostate | 0.660 | 0.005 | 0.658 | 0.013 |
| | Testis | 0.740 | 0.096 | 0.737 | 0.000 |
| | Kidney | 0.673 | 0.014 | 0.669 | 0.001 |
| | Bladder | 0.758 | 0.009 | 0.755 | 0.002 |
| | Brain | 0.587 | 0.019 | 0.584 | 0.001 |
| | Thyroid | 0.660 | 0.027 | 0.661 | 0.000 |
| | Non-Hodgkin Lymphoma | 0.643 | 0.012 | 0.639 | 0.001 |
| | Multiple Myeloma | 0.671 | 0.017 | 0.668 | 0.001 |
| | AML | 0.709 | 0.038 | 0.706 | 0.000 |
| | Other | 0.650 | 0.003 | 0.648 | 0.015 |
| | Death | 0.791 | 0.004 | 0.790 | 0.009 |

#### **ST3:** Model comparison.

| | Cancer | Log-Likelihood | | | | | P-Values (FWER-Corrected) | | | | | |
|---|---|---|---|---|---|---|---|---|---|---|---|---|
| | | Base | Base + Health | Base + Gene | All | UKB-AgeSex | Base / All | Base / Health | Base / Gene | Base+Health / All | Base+Gene / All | UKB-AgeSex / All |
| Denmark | Oesophagus | -20326.662 | -20097.864 | -20321.159 | -20093.123 | ·· | 0.000 | 0.000 | 0.021 | 0.046 | 0.000 | ·· |
| | Stomach | -22039.178 | -21991.113 | -22033.892 | -21985.542 | ·· | 0.000 | 0.000 | 0.025 | 0.019 | 0.000 | ·· |
| | Colorectal | -153697.350 | -153600.420 | -153620.910 | -153528.560 | ·· | 0.000 | 0.000 | 0.000 | 0.000 | 0.000 | ·· |
| | Liver | -16547.177 | -15631.003 | -16546.814 | -15629.177 | ·· | 0.000 | 0.000 | 0.901 | 0.560 | 0.000 | ·· |
| | Pancreas | -30025.035 | -29888.958 | -30024.890 | -29888.938 | ·· | 0.000 | 0.000 | 0.901 | 0.901 | 0.000 | ·· |
| | Lung | -131119.800 | -129100.720 | -131063.150 | -129041.780 | ·· | 0.000 | 0.000 | 0.000 | 0.000 | 0.000 | ·· |
| | Melanoma | -82073.718 | -81856.785 | -81909.540 | -81708.856 | ·· | 0.000 | 0.000 | 0.000 | 0.000 | 0.000 | ·· |
| | Breast | -160303.310 | -160080.800 | -160140.350 | -159937.230 | ·· | 0.000 | 0.000 | 0.000 | 0.000 | 0.000 | ·· |
| | Cervix Uteri | -14084.641 | -14029.227 | -14052.087 | -13996.080 | ·· | 0.000 | 0.000 | 0.000 | 0.000 | 0.000 | ·· |
| | Corpus Uteri | -24088.914 | -23882.060 | -24087.109 | -23881.203 | ·· | 0.000 | 0.000 | 0.574 | 0.901 | 0.000 | ·· |
| | Ovary | -17526.526 | -17523.697 | -17524.498 | -17522.032 | ·· | 0.179 | 0.238 | 0.485 | 0.635 | 0.316 | ·· |
| | Prostate | -128125.440 | -127932.410 | -127910.540 | -127718.150 | ·· | 0.000 | 0.000 | 0.000 | 0.000 | 0.000 | ·· |
| | Testis | -14680.359 | -14661.843 | -14586.911 | -14577.004 | ·· | 0.000 | 0.000 | 0.000 | 0.000 | 0.000 | ·· |
| | Kidney | -31369.039 | -31221.597 | -31369.031 | -31221.459 | ·· | 0.000 | 0.000 | 0.901 | 0.901 | 0.000 | ·· |
| | Bladder | -24361.172 | -24232.906 | -24339.634 | -24210.174 | ·· | 0.000 | 0.000 | 0.000 | 0.000 | 0.000 | ·· |
| | Brain | -19830.430 | -19707.512 | -19828.016 | -19705.472 | ·· | 0.000 | 0.000 | 0.336 | 0.477 | 0.000 | ·· |
| | Thyroid | -15117.174 | -15010.054 | -15104.929 | -14999.389 | ·· | 0.000 | 0.000 | 0.000 | 0.000 | 0.000 | ·· |
| | Non-Hodgkin Lymphoma | -38351.686 | -38285.587 | -38350.698 | -38284.298 | ·· | 0.000 | 0.000 | 0.901 | 0.867 | 0.000 | ·· |
| | Multiple Myeloma | -14756.177 | -14663.027 | -14756.030 | -14662.980 | ·· | 0.000 | 0.000 | 0.901 | 0.901 | 0.000 | ·· |
| | AML | -6470.858 | -6395.330 | -6468.353 | -6393.188 | ·· | 0.000 | 0.000 | 0.302 | 0.423 | 0.000 | ·· |
| | Other | -415320.660 | -414354.960 | -415245.180 | -414287.250 | ·· | 0.000 | 0.000 | 0.000 | 0.000 | 0.000 | ·· |
| | Death | -542139.070 | -521669.520 | -542094.930 | -521584.520 | ·· | 0.000 | 0.000 | 0.000 | 0.000 | 0.000 | ·· |
| UK Biobank | Oesophagus | -3531.5363 | -3503.0347 | -3530.8549 | -3502.4515 | -3520.8277 | 0.000 | 0.000 | 1.000 | 1.000 | 0.000 | 0.000 |
| | Stomach | -2768.6624 | -2758.5274 | -2768.4287 | -2758.3464 | -2765.9961 | 0.002 | 0.000 | 1.000 | 1.000 | 0.001 | 0.027 |
| | Colorectal | -18342.54 | -18307.128 | -18330.719 | -18294.988 | -18338.608 | 0.000 | 0.000 | 0.000 | 0.000 | 0.000 | 0.000 |
| | Liver | -2374.805 | -2321.8366 | -2373.8003 | -2320.9145 | -2370.1542 | 0.000 | 0.000 | 1.000 | 1.000 | 0.000 | 0.000 |
| | Pancreas | -4453.0337 | -4446.6233 | -4452.7434 | -4446.2871 | -4450.3698 | 0.060 | 0.020 | 1.000 | 1.000 | 0.019 | 0.691 |
| | Lung | -13613.933 | -13218.56 | -13585.766 | -13197.524 | -13580.378 | 0.000 | 0.000 | 0.000 | 0.000 | 0.000 | 0.000 |
| | Melanoma | -8329.6242 | -8325.2196 | -8329.5261 | -8325.1243 | -8316.4255 | 0.522 | 0.147 | 1.000 | 1.000 | 0.147 | 1.000 |
| | Breast | -26872.947 | -26840.228 | -26839.827 | -26808.835 | -26866.537 | 0.000 | 0.000 | 0.000 | 0.000 | 0.000 | 0.000 |
| | Cervix Uteri | -390.77261 | -389.71526 | -390.211 | -389.1597 | -390.68294 | 1.000 | 1.000 | 1.000 | 1.000 | 1.000 | 1.000 |
| | Corpus Uteri | -4524.5095 | -4486.6616 | -4524.3545 | -4486.4278 | -4524.2196 | 0.000 | 0.000 | 1.000 | 1.000 | 0.000 | 0.000 |
| | Ovary | -3227.4494 | -3226.7124 | -3226.365 | -3225.6125 | -3225.0276 | 1.000 | 1.000 | 1.000 | 1.000 | 1.000 | 1.000 |
| | Prostate | -26920.688 | -26910.062 | -26893.111 | -26882.965 | -26896.436 | 0.000 | 0.000 | 0.000 | 0.000 | 0.000 | 0.000 |
| | Testis | -132.0101 | -126.92288 | -130.50462 | -125.3686 | -132.09463 | 0.067 | 0.073 | 1.000 | 1.000 | 0.069 | 0.061 |
| | Kidney | -4532.2661 | -4526.9541 | -4532.1961 | -4526.8798 | -4528.2026 | 0.224 | 0.058 | 1.000 | 1.000 | 0.058 | 1.000 |
| | Bladder | -7577.9878 | -7559.3102 | -7573.8966 | -7555.19 | -7562.0885 | 0.000 | 0.000 | 0.207 | 0.201 | 0.000 | 0.052 |
| | Brain | -2667.9257 | -2667.5505 | -2667.5118 | -2667.1313 | -2664.0967 | 1.000 | 1.000 | 1.000 | 1.000 | 1.000 | 1.000 |
| | Thyroid | -1196.4797 | -1180.7691 | -1195.4041 | -1179.6977 | -1187.8032 | 0.000 | 0.000 | 1.000 | 1.000 | 0.000 | 0.018 |
| | Non-Hodgkin Lymphoma | -6684.3819 | -6670.0451 | -6684.3571 | -6670.0311 | -6678.2122 | 0.000 | 0.000 | 1.000 | 1.000 | 0.000 | 0.017 |
| | Multiple Myeloma | -3113.8261 | -3103.4745 | -3113.8099 | -3103.4495 | -3109.174 | 0.002 | 0.000 | 1.000 | 1.000 | 0.000 | 0.160 |
| | AML | -711.75173 | -684.64681 | -711.72603 | -684.60737 | -710.47767 | 0.000 | 0.000 | 1.000 | 1.000 | 0.000 | 0.000 |
| | Other | -75270.965 | -75222.28 | -75268.819 | -75220.573 | -75232.146 | 0.000 | 0.000 | 0.980 | 1.000 | 0.000 | 0.001 |
| | Death | -42397.76 | -40780.501 | -42385.376 | -40774.579 | -42238.397 | 0.000 | 0.000 | 0.000 | 0.032 | 0.000 | 0.000 |

**Notes:** Base := Breslow Estimate for the baseline hazard based on the Danish data, Health := Diseases + Basic health factors, UKB-AgeSex := Age-Sex model fitted on the UKB data.
/ Indicates the different models that are tested against each other. FWER := Family-wise Error Rate

**ST4:** Kaplan Meier curve comparison of top risk quantiles between our predictions and age-sex alone.

| | Denmark | | | | | | UK Biobank | | | | | |
|---|---|---|---|---|---|---|---|---|---|---|---|---|
| **Cancer** | Hazard Ratio (HR) | HR - lower | HR - upper | LogRank Statistic | P-Value | P-Value (FWER) | Hazard Ratio (HR) | HR - lower | HR - upper | LogRank Statistic | P-Value | P-Value (FWER) |
| Oesophagus | 1.974 | 1.566 | 2.489 | 34.435 | 0.000 | 0.000 | 2.623 | 1.096 | 6.281 | 5.063 | 0.024 | 0.391 |
| Stomach | 1.127 | 0.857 | 1.482 | 0.738 | 0.390 | 0.648 | 1.691 | 0.615 | 4.654 | 1.060 | 0.303 | 0.861 |
| Colorectal | 0.975 | 0.874 | 1.088 | 0.208 | 0.648 | 0.648 | 1.171 | 0.729 | 1.879 | 0.427 | 0.513 | 0.861 |
| Liver | 4.116 | 3.283 | 5.159 | 177.442 | 0.000 | 0.000 | 3.589 | 1.636 | 7.875 | 11.623 | 0.001 | 0.012 |
| Pancreas | 1.659 | 1.314 | 2.095 | 18.509 | 0.000 | 0.000 | 1.534 | 0.627 | 3.752 | 0.891 | 0.345 | 0.861 |
| Lung | 2.404 | 2.175 | 2.658 | 314.215 | 0.000 | 0.000 | 4.266 | 2.800 | 6.499 | 54.207 | 0.000 | 0.000 |
| Melanoma | 1.555 | 1.262 | 1.916 | 17.478 | 0.000 | 0.000 | 0.922 | 0.445 | 1.911 | 0.047 | 0.828 | 0.861 |
| Breast | 1.326 | 1.139 | 1.544 | 13.345 | 0.000 | 0.003 | 1.945 | 1.235 | 3.064 | 8.550 | 0.003 | 0.062 |
| Cervix Uteri | 1.196 | 0.589 | 2.426 | 0.246 | 0.620 | 0.648 | .. | .. | .. | .. | .. | .. |
| Corpus Uteri | 2.615 | 1.941 | 3.521 | 43.207 | 0.000 | 0.000 | 6.895 | 2.385 | 19.938 | 17.174 | 0.000 | 0.001 |
| Ovary | 1.369 | 0.915 | 2.048 | 2.354 | 0.125 | 0.625 | 3.316 | 0.345 | 31.883 | 1.213 | 0.271 | 0.861 |
| Prostate | 1.255 | 1.117 | 1.409 | 14.803 | 0.000 | 0.001 | 1.287 | 0.854 | 1.938 | 1.463 | 0.226 | 0.861 |
| Testis | 1.909 | 1.144 | 3.185 | 6.349 | 0.012 | 0.081 | .. | .. | .. | .. | .. | .. |
| Kidney | 1.567 | 1.212 | 2.025 | 11.978 | 0.001 | 0.005 | 1.253 | 0.519 | 3.024 | 0.253 | 0.615 | 0.861 |
| Bladder | 1.659 | 1.342 | 2.051 | 22.336 | 0.000 | 0.000 | 1.110 | 0.687 | 1.793 | 0.183 | 0.669 | 0.861 |
| Brain | 2.446 | 1.643 | 3.641 | 20.764 | 0.000 | 0.000 | 1.427 | 0.453 | 4.497 | 0.373 | 0.541 | 0.861 |
| Thyroid | 5.063 | 2.965 | 8.645 | 43.730 | 0.000 | 0.000 | 3.814 | 1.050 | 13.858 | 4.791 | 0.029 | 0.435 |
| Non-Hodgkin Lymphoma | 1.124 | 0.885 | 1.426 | 0.918 | 0.338 | 0.648 | 1.802 | 0.796 | 4.078 | 2.056 | 0.152 | 0.861 |
| Multiple Myeloma | 1.827 | 1.285 | 2.596 | 11.618 | 0.001 | 0.006 | 1.452 | 0.553 | 3.814 | 0.579 | 0.447 | 0.861 |
| AML | 1.585 | 1.062 | 2.366 | 5.166 | 0.023 | 0.138 | 4.068 | 0.864 | 19.156 | 3.702 | 0.054 | 0.707 |
| Other | 1.090 | 1.013 | 1.171 | 5.396 | 0.020 | 0.121 | 0.980 | 0.782 | 1.228 | 0.030 | 0.861 | 0.861 |
| Death | 3.292 | 3.156 | 3.433 | 3468.066 | 0.000 | 0.000 | 4.618 | 3.841 | 5.552 | 320.842 | 0.000 | 0.000 |

**ST5:** Performance comparison to QCancer

| Cancer | Ours - ROC-AUC | QCancer - ROC-AUC | |
|---|---|---|---|
| | | Female | Male |
| Oesophagus | 0.889 | 0.863 | 0.856 |
| Stomach | 0.852 | .. | .. |
| Colorectal | 0.845 | 0.841 | 0.851 |
| Liver | 0.903 | .. | .. |
| Pancreas | 0.857 | 0.865 | 0.851 |
| Lung | 0.885 | 0.9 | 0.901 |
| Melanoma | 0.703 | .. | .. |
| Breast | 0.753 | 0.747 | .. |
| Cervix Uteri | 0.659 | .. | .. |
| Corpus Uteri | 0.849 | 0.818 | .. |
| Ovary | 0.786 | 0.764 | .. |
| Prostate | 0.890 | .. | 0.794 |
| Testis | 0.691 | .. | .. |
| Kidney | 0.827 | 0.849 | 0.856 |
| Bladder | 0.890 | .. | .. |
| Brain | 0.751 | .. | .. |
| Thyroid | 0.718 | .. | .. |
| Non-Hodgkin Lymphoma | 0.793 | .. | .. |
| Multiple Myeloma | 0.846 | .. | .. |
| AML | 0.797 | 0.798 | 0.79 |
| Other | 0.796 | .. | .. |
| Death | 0.887 | .. | .. |

### **ST6:** Screening evaluation

| | Cancer | Age | Baseline | | | | | | | | | Population health informed screening | | | | | | | | |
|---|---|---|---|---|---|---|---|---|---|---|---|---|---|---|---|---|---|---|---|---|
| | | | N | Cancers | Age - mean | Age - s.d. | Age - q10 | Age - q25 | Age - median | Age - q75 | Age - q90 | N | Cancers | Age - mean | Age - s.d. | Age - q10 | Age - q25 | Age - median | Age - q75 | Age - q90 |
| Denmark | Oesophagus | 40 | 397,561 | 14 | 43.34 | 1.29 | 41.73 | 42.53 | 43.90 | 44.14 | 44.55 | 397,561 | 17 | 42.63 | 1.92 | 39.47 | 41.42 | 43.81 | 43.91 | 44.45 |
| | Oesophagus | 45 | 411,925 | 60 | 47.51 | 1.56 | 45.52 | 46.15 | 47.33 | 49.08 | 49.55 | 411,926 | 61 | 47.27 | 1.84 | 45.03 | 45.76 | 47.22 | 49.07 | 49.55 |
| | Oesophagus | 50 | 382,840 | 125 | 52.83 | 1.36 | 50.82 | 51.82 | 52.91 | 53.95 | 54.58 | 382,840 | 133 | 52.05 | 2.44 | 48.96 | 50.98 | 52.68 | 53.83 | 54.54 |
| | Oesophagus | 55 | 332,946 | 240 | 57.82 | 1.42 | 55.76 | 56.65 | 57.95 | 59.16 | 59.69 | 332,946 | 275 | 56.44 | 3.18 | 52.35 | 54.68 | 57.46 | 58.78 | 59.57 |
| | Oesophagus | 60 | 299,441 | 326 | 62.72 | 1.40 | 60.69 | 61.65 | 62.88 | 63.94 | 64.49 | 299,441 | 403 | 60.39 | 3.98 | 54.64 | 58.24 | 61.63 | 63.50 | 64.34 |
| | Oesophagus | 65 | 292,150 | 351 | 67.72 | 1.41 | 65.76 | 66.52 | 67.75 | 68.96 | 69.58 | 292,150 | 486 | 64.15 | 4.85 | 57.03 | 61.22 | 65.39 | 68.11 | 69.15 |
| | Stomach | 40 | 397,561 | 37 | 42.85 | 1.43 | 40.69 | 41.67 | 43.32 | 43.90 | 44.60 | 397,561 | 42 | 42.40 | 1.82 | 39.52 | 40.98 | 43.13 | 43.84 | 44.53 |
| | Stomach | 45 | 411,925 | 104 | 47.72 | 1.47 | 45.63 | 46.46 | 47.81 | 48.96 | 49.60 | 411,925 | 100 | 47.39 | 2.09 | 44.66 | 46.15 | 47.81 | 48.96 | 49.61 |
| | Stomach | 50 | 382,840 | 136 | 52.80 | 1.37 | 50.83 | 51.83 | 52.93 | 53.86 | 54.55 | 382,840 | 151 | 52.15 | 2.12 | 49.12 | 50.89 | 52.64 | 53.71 | 54.51 |
| | Stomach | 55 | 332,946 | 224 | 57.69 | 1.36 | 55.68 | 56.65 | 57.66 | 58.84 | 59.53 | 332,946 | 235 | 56.94 | 2.44 | 53.45 | 55.88 | 57.47 | 58.79 | 59.49 |
| | Stomach | 60 | 299,441 | 297 | 62.60 | 1.47 | 60.43 | 61.35 | 62.71 | 63.94 | 64.47 | 299,441 | 317 | 61.87 | 2.43 | 58.47 | 60.37 | 62.41 | 63.83 | 64.43 |
| | Stomach | 65 | 292,150 | 385 | 67.70 | 1.43 | 65.78 | 66.47 | 67.72 | 68.95 | 69.62 | 292,150 | 415 | 66.64 | 2.75 | 62.89 | 65.15 | 67.16 | 68.86 | 69.55 |
| | Colorectal | 40 | 397,561 | 220 | 42.77 | 1.32 | 40.87 | 41.70 | 42.88 | 43.85 | 44.46 | 397,561 | 236 | 42.09 | 2.55 | 38.98 | 41.35 | 42.72 | 43.75 | 44.45 |
| | Colorectal | 45 | 411,925 | 574 | 47.83 | 1.34 | 45.80 | 46.87 | 47.89 | 48.94 | 49.59 | 411,934 | 580 | 47.52 | 2.01 | 45.33 | 46.61 | 47.84 | 48.91 | 49.57 |
| | Colorectal | 50 | 382,840 | 871 | 52.72 | 1.38 | 50.78 | 51.62 | 52.79 | 53.90 | 54.56 | 382,840 | 937 | 52.15 | 2.18 | 49.12 | 51.12 | 52.54 | 53.83 | 54.50 |
| | Colorectal | 55 | 332,946 | 1,316 | 57.63 | 1.46 | 55.54 | 56.35 | 57.69 | 58.89 | 59.57 | 332,947 | 1,362 | 56.95 | 2.50 | 53.68 | 55.77 | 57.43 | 58.83 | 59.56 |
| | Colorectal | 60 | 299,441 | 1,997 | 62.66 | 1.43 | 60.64 | 61.45 | 62.75 | 63.91 | 64.57 | 299,441 | 1,988 | 61.47 | 3.08 | 57.11 | 60.00 | 62.31 | 63.84 | 64.55 |
| | Colorectal | 65 | 292,150 | 2,711 | 67.66 | 1.42 | 65.62 | 66.51 | 67.75 | 68.87 | 69.61 | 292,150 | 2,688 | 66.31 | 3.51 | 60.94 | 65.00 | 67.47 | 68.77 | 69.55 |
| | Liver | 40 | 397,561 | 15 | 42.88 | 1.56 | 41.08 | 41.66 | 42.79 | 44.42 | 44.86 | 397,563 | 16 | 42.35 | 2.53 | 40.50 | 41.51 | 42.62 | 44.32 | 44.86 |
| | Liver | 45 | 411,925 | 33 | 47.88 | 1.46 | 45.97 | 46.69 | 48.08 | 49.27 | 49.76 | 411,925 | 39 | 46.82 | 2.86 | 44.15 | 45.72 | 47.32 | 49.09 | 49.53 |
| | Liver | 50 | 382,840 | 94 | 53.02 | 1.36 | 51.18 | 51.94 | 53.03 | 54.29 | 54.76 | 382,840 | 112 | 51.86 | 3.16 | 47.53 | 50.93 | 52.70 | 54.09 | 54.73 |
| | Liver | 55 | 332,946 | 173 | 57.71 | 1.43 | 55.75 | 56.57 | 57.76 | 59.11 | 59.50 | 332,946 | 237 | 55.92 | 3.37 | 51.53 | 54.25 | 56.77 | 58.42 | 59.45 |
| | Liver | 60 | 299,441 | 235 | 62.61 | 1.46 | 60.73 | 61.21 | 62.78 | 63.80 | 64.66 | 299,441 | 374 | 59.64 | 4.22 | 53.67 | 57.27 | 60.60 | 63.03 | 64.33 |
| | Liver | 65 | 292,150 | 281 | 67.66 | 1.45 | 65.54 | 66.52 | 67.72 | 68.96 | 69.52 | 292,150 | 517 | 63.17 | 5.44 | 55.54 | 59.57 | 64.48 | 67.68 | 69.14 |
| | Pancreas | 40 | 397,561 | 39 | 42.97 | 1.26 | 41.10 | 42.03 | 43.27 | 44.09 | 44.52 | 397,561 | 38 | 42.72 | 2.05 | 41.06 | 41.95 | 43.28 | 44.11 | 44.53 |
| | Pancreas | 45 | 411,925 | 103 | 47.91 | 1.30 | 46.28 | 46.83 | 47.91 | 49.14 | 49.68 | 411,925 | 102 | 47.85 | 1.43 | 46.28 | 46.81 | 47.94 | 49.19 | 49.69 |
| | Pancreas | 50 | 382,840 | 175 | 52.67 | 1.39 | 50.75 | 51.65 | 52.64 | 54.00 | 54.64 | 382,840 | 187 | 52.02 | 2.26 | 48.26 | 50.95 | 52.40 | 53.81 | 54.60 |
| | Pancreas | 55 | 332,946 | 268 | 57.81 | 1.42 | 55.73 | 56.76 | 57.76 | 59.13 | 59.66 | 332,946 | 300 | 56.78 | 2.77 | 53.17 | 55.16 | 57.44 | 58.98 | 59.59 |
| | Pancreas | 60 | 299,441 | 370 | 62.68 | 1.39 | 60.74 | 61.50 | 62.80 | 63.87 | 64.57 | 299,441 | 402 | 61.41 | 3.15 | 57.55 | 59.91 | 62.16 | 63.76 | 64.54 |
| | Pancreas | 65 | 292,150 | 561 | 67.59 | 1.44 | 65.48 | 66.40 | 67.68 | 68.81 | 69.53 | 292,150 | 584 | 66.47 | 3.16 | 63.26 | 65.18 | 67.07 | 68.59 | 69.43 |
| | Lung | 40 | 397,561 | 86 | 43.00 | 1.40 | 40.74 | 42.11 | 43.30 | 44.03 | 44.76 | 397,561 | 95 | 42.30 | 2.22 | 39.19 | 40.64 | 43.16 | 43.99 | 44.71 |
| | Lung | 45 | 411,925 | 300 | 48.06 | 1.39 | 45.96 | 47.04 | 48.32 | 49.26 | 49.73 | 411,933 | 307 | 47.45 | 2.38 | 44.32 | 46.41 | 48.14 | 49.16 | 49.73 |
| | Lung | 50 | 382,840 | 713 | 52.75 | 1.42 | 50.68 | 51.53 | 52.91 | 53.97 | 54.57 | 382,840 | 785 | 51.67 | 2.68 | 48.08 | 50.03 | 52.42 | 53.83 | 54.52 |
| | Lung | 55 | 332,946 | 1,195 | 57.74 | 1.46 | 55.61 | 56.50 | 57.91 | 58.95 | 59.64 | 332,959 | 1,412 | 55.93 | 3.42 | 50.90 | 53.92 | 56.82 | 58.68 | 59.52 |
| | Lung | 60 | 299,441 | 1,861 | 62.64 | 1.43 | 60.64 | 61.44 | 62.72 | 63.88 | 64.54 | 299,441 | 2,243 | 60.85 | 3.48 | 56.07 | 59.06 | 61.70 | 63.52 | 64.42 |
| | Lung | 65 | 292,150 | 2,445 | 67.60 | 1.42 | 65.60 | 66.37 | 67.68 | 68.80 | 69.54 | 292,150 | 3,152 | 65.40 | 3.72 | 60.33 | 63.60 | 66.24 | 68.21 | 69.29 |
| | Breast | 40 | 192,944 | 886 | 42.77 | 1.40 | 40.68 | 41.67 | 42.82 | 43.99 | 44.66 | 192,944 | 916 | 42.44 | 1.94 | 39.80 | 41.41 | 42.77 | 43.91 | 44.65 |
| | Breast | 45 | 198,309 | 1,442 | 47.86 | 1.40 | 45.72 | 46.74 | 48.06 | 49.06 | 49.63 | 198,309 | 1,513 | 47.18 | 2.32 | 43.89 | 46.01 | 47.72 | 48.96 | 49.58 |
| | Breast | 50 | 184,578 | 1,561 | 52.32 | 1.46 | 50.43 | 51.05 | 52.13 | 53.63 | 54.45 | 184,578 | 1,709 | 50.41 | 3.20 | 46.05 | 48.49 | 50.75 | 53.08 | 54.32 |
| | Breast | 55 | 161,464 | 1,476 | 57.54 | 1.45 | 55.54 | 56.25 | 57.54 | 58.80 | 59.55 | 161,464 | 1,749 | 54.22 | 4.36 | 47.96 | 50.90 | 55.22 | 57.93 | 59.26 |
| | Breast | 60 | 147,139 | 1,880 | 62.48 | 1.45 | 60.48 | 61.23 | 62.45 | 63.71 | 64.53 | 147,139 | 1,981 | 57.72 | 5.31 | 49.55 | 53.98 | 58.98 | 62.10 | 63.76 |
| | Breast | 65 | 145,266 | 1,924 | 67.34 | 1.40 | 65.48 | 66.15 | 67.28 | 68.51 | 69.35 | 145,266 | 2,140 | 59.52 | 6.09 | 50.34 | 55.32 | 60.55 | 64.27 | 66.91 |
| | Corpus Uteri | 40 | 192,944 | 40 | 42.90 | 1.33 | 41.19 | 41.80 | 42.93 | 43.86 | 44.77 | 192,944 | 51 | 41.42 | 2.81 | 37.56 | 39.60 | 41.86 | 43.64 | 44.58 |
| | Corpus Uteri | 45 | 198,309 | 99 | 47.73 | 1.46 | 45.88 | 46.52 | 47.63 | 49.20 | 49.71 | 198,309 | 108 | 46.95 | 2.39 | 43.72 | 45.92 | 47.31 | 48.86 | 49.67 |
| | Corpus Uteri | 50 | 184,578 | 190 | 52.66 | 1.43 | 50.61 | 51.49 | 52.88 | 53.89 | 54.56 | 184,578 | 209 | 51.50 | 2.72 | 47.76 | 49.95 | 51.97 | 53.56 | 54.45 |
| | Corpus Uteri | 55 | 161,464 | 272 | 57.56 | 1.45 | 55.49 | 56.32 | 57.72 | 58.73 | 59.51 | 161,464 | 331 | 55.77 | 3.26 | 51.25 | 53.59 | 56.67 | 58.35 | 59.30 |
| | Corpus Uteri | 60 | 147,139 | 326 | 62.59 | 1.39 | 60.58 | 61.38 | 62.62 | 63.77 | 64.52 | 147,139 | 423 | 59.85 | 3.99 | 53.95 | 57.66 | 60.76 | 63.02 | 64.15 |
| | Corpus Uteri | 65 | 145,266 | 424 | 67.67 | 1.44 | 65.65 | 66.45 | 67.71 | 68.93 | 69.60 | 145,266 | 538 | 64.29 | 4.77 | 57.27 | 61.70 | 65.68 | 68.07 | 69.26 |
| | Ovary | 40 | 192,944 | 68 | 42.68 | 1.50 | 40.73 | 41.39 | 42.71 | 44.02 | 44.62 | 192,944 | 71 | 41.87 | 2.46 | 38.10 | 40.63 | 42.18 | 43.88 | 44.61 |
| | Ovary | 45 | 198,309 | 105 | 47.89 | 1.48 | 45.63 | 46.74 | 48.09 | 49.06 | 49.79 | 198,309 | 113 | 46.69 | 2.74 | 42.09 | 44.84 | 47.83 | 48.75 | 49.73 |
| | Ovary | 50 | 184,578 | 119 | 52.66 | 1.42 | 50.56 | 51.57 | 52.75 | 53.92 | 54.50 | 184,578 | 133 | 50.60 | 3.06 | 46.63 | 48.42 | 51.16 | 53.05 | 54.10 |
| | Ovary | 55 | 161,464 | 164 | 57.57 | 1.44 | 55.56 | 56.22 | 57.59 | 58.74 | 59.46 | 161,464 | 164 | 55.29 | 3.81 | 49.15 | 52.74 | 56.18 | 58.35 | 59.37 |
| | Ovary | 60 | 147,139 | 202 | 62.56 | 1.45 | 60.56 | 61.35 | 62.66 | 63.80 | 64.48 | 147,139 | 205 | 60.48 | 4.10 | 54.63 | 58.67 | 61.84 | 63.41 | 64.36 |
| | Ovary | 65 | 145,266 | 269 | 67.57 | 1.45 | 65.58 | 66.43 | 67.64 | 68.80 | 69.54 | 145,266 | 274 | 66.11 | 3.32 | 61.85 | 64.80 | 66.88 | 68.56 | 69.35 |
| | Prostate | 40 | 204,617 | 30 | 42.88 | 1.55 | 40.40 | 41.74 | 43.37 | 44.07 | 44.63 | 204,617 | 30 | 42.83 | 1.66 | 40.40 | 41.74 | 43.37 | 44.07 | 44.63 |
| | Prostate | 45 | 213,616 | 163 | 48.09 | 1.28 | 45.91 | 47.42 | 48.34 | 49.05 | 49.64 | 213,619 | 168 | 47.92 | 1.57 | 45.71 | 47.13 | 48.33 | 49.00 | 49.64 |
| | Prostate | 50 | 198,262 | 572 | 52.92 | 1.39 | 50.83 | 51.85 | 53.19 | 54.14 | 54.59 | 198,262 | 590 | 52.71 | 1.71 | 50.34 | 51.69 | 53.09 | 54.10 | 54.59 |
| | Prostate | 55 | 171,482 | 1,174 | 57.80 | 1.44 | 55.67 | 56.67 | 57.90 | 59.08 | 59.67 | 171,482 | 1,246 | 57.39 | 2.03 | 54.55 | 56.31 | 57.80 | 59.02 | 59.66 |
| | Prostate | 60 | 152,302 | 2,030 | 62.70 | 1.44 | 60.63 | 61.50 | 62.78 | 63.95 | 64.66 | 152,302 | 2,198 | 62.02 | 2.38 | 58.92 | 60.76 | 62.56 | 63.85 | 64.63 |
| | Prostate | 65 | 146,884 | 3,016 | 67.63 | 1.41 | 65.61 | 66.43 | 67.68 | 68.84 | 69.54 | 146,884 | 3,224 | 66.43 | 2.92 | 62.26 | 64.92 | 67.18 | 68.65 | 69.48 |
| | Kidney | 40 | 397,561 | 81 | 42.62 | 1.50 | 40.60 | 41.35 | 42.77 | 43.89 | 44.66 | 397,561 | 92 | 41.88 | 2.49 | 38.90 | 40.60 | 42.29 | 43.80 | 44.64 |
| | Kidney | 45 | 411,925 | 174 | 47.79 | 1.41 | 45.72 | 46.69 | 47.85 | 49.05 | 49.63 | 411,930 | 193 | 46.88 | 2.98 | 42.90 | 46.08 | 47.62 | 48.83 | 49.62 |
| | Kidney | 50 | 382,840 | 241 | 52.61 | 1.41 | 50.69 | 51.44 | 52.59 | 53.92 | 54.48 | 382,840 | 280 | 51.42 | 3.10 | 47.84 | 49.82 | 52.12 | 53.70 | 54.44 |
| | Kidney | 55 | 332,946 | 325 | 57.44 | 1.45 | 55.56 | 56.14 | 57.38 | 58.63 | 59.55 | 332,946 | 365 | 56.19 | 2.98 | 52.01 | 54.65 | 56.76 | 58.45 | 59.51 |
| | Kidney | 60 | 299,441 | 396 | 62.51 | 1.46 | 60.54 | 61.16 | 62.50 | 63.76 | 64.56 | 299,441 | 445 | 60.82 | 3.56 | 55.83 | 59.08 | 61.77 | 63.62 | 64.49 |
| | Kidney | 65 | 292,150 | 471 | 67.67 | 1.40 | 65.73 | 66.54 | 67.67 | 68.87 | 69.53 | 292,150 | 539 | 65.32 | 4.06 | 59.67 | 63.29 | 66.47 | 68.51 | 69.43 |
| | Bladder | 40 | 397,561 | 18 | 42.82 | 1.53 | 40.62 | 41.54 | 42.88 | 44.34 | 44.60 | 397,561 | 20 | 42.20 | 2.23 | 39.39 | 40.64 | 42.63 | 44.32 | 44.60 |
| | Bladder | 45 | 411,925 | 53 | 47.86 | 1.39 | 45.65 | 46.72 | 48.12 | 48.82 | 49.75 | 411,925 | 58 | 47.34 | 2.12 | 44.63 | 46.21 | 47.95 | 48.70 | 49.68 |
| | Bladder | 50 | 382,840 | 108 | 52.58 | 1.43 | 50.48 | 51.46 | 52.74 | 53.88 | 54.48 | 382,840 | 111 | 52.11 | 2.41 | 49.36 | 51.24 | 52.53 | 53.88 | 54.47 |
| | Bladder | 55 | 332,946 | 187 | 57.82 | 1.47 | 55.62 | 56.62 | 58.03 | 59.15 | 59.63 | 332,946 | 196 | 57.31 | 2.50 | 54.52 | 56.23 | 57.96 | 59.12 | 59.62 |
| | Bladder | 60 | 299,441 | 336 | 62.64 | 1.40 | 60.65 | 61.46 | 62.76 | 63.84 | 64.56 | 299,441 | 350 | 62.15 | 2.21 | 59.25 | 61.12 | 62.58 | 63.78 | 64.55 |
| | Bladder | 65 | 292,150 | 479 | 67.69 | 1.38 | 65.72 | 66.48 | 67.83 | 68.89 | 69.51 | 292,150 | 500 | 67.01 | 2.46 | 64.11 | 65.89 | 67.62 | 68.84 | 69.46 |
| | Brain | 40 | 397,561 | 74 | 42.80 | 1.39 | 40.50 | 41.90 | 42.82 | 43.98 | 44.55 | 397,561 | 95 | 40.35 | 5.27 | 32.73 | 39.67 | 42.47 | 43.63 | 44.45 |
| | Brain | 45 | 411,925 | 121 | 47.58 | 1.36 | 45.71 | 46.60 | 47.43 | 48.79 | 49.53 | 411,925 | 138 | 45.14 | 5.64 | 37.92 | 44.35 | 47.02 | 48.59 | 49.42 |
| | Brain | 50 | 382,840 | 135 | 52.49 | 1.40 | 50.49 | 51.30 | 52.57 | 53.71 | 54.38 | 382,840 | 165 | 49.32 | 6.69 | 40.11 | 48.28 | 51.92 | 53.25 | 54.33 |
| | Brain | 55 | 332,946 | 171 | 57.59 | 1.36 | 55.63 | 56.45 | 57.55 | 58.78 | 59.57 | 332,946 | 204 | 53.89 | 7.67 | 43.83 | 53.13 | 56.92 | 58.22 | 59.35 |
| | Brain | 60 | 299,441 | 207 | 62.68 | 1.43 | 60.58 | 61.53 | 62.72 | 63.88 | 64.62 | 299,441 | 242 | 58.82 | 8.21 | 46.93 | 58.87 | 62.10 | 63.68 | 64.58 |
| | Brain | 65 | 292,150 | 242 | 67.71 | 1.51 | 65.66 | 66.32 | 67.74 | 69.09 | 69.74 | 292,150 | 289 | 63.35 | 8.83 | 51.06 | 63.38 | 66.32 | 68.60 | 69.60 |
| | NHL | 40 | 397,561 | 112 | 42.73 | 1.42 | 40.69 | 41.62 | 42.65 | 44.01 | 44.56 | 397,561 | 127 | 41.53 | 3.45 | 37.47 | 40.38 | 42.41 | 43.81 | 44.49 |
| | NHL | 45 | 411,925 | 173 | 47.92 | 1.33 | 46.02 | 46.78 | 48.07 | 49.11 | 49.57 | 411,925 | 184 | 46.97 | 3.21 | 43.95 | 46.06 | 47.87 | 49.00 | 49.53 |
| | NHL | 50 | 382,840 | 234 | 52.62 | 1.45 | 50.68 | 51.35 | 52.69 | 53.93 | 54.65 | 382,849 | 253 | 51.50 | 3.17 | 47.93 | 50.03 | 52.28 | 53.80 | 54.60 |
| | NHL | 55 | 332,946 | 358 | 57.55 | 1.44 | 55.54 | 56.32 | 57.52 | 58.74 | 59.51 | 332,946 | 356 | 56.35 | 3.57 | 51.92 | 55.10 | 57.43 | 58.72 | 59.50 |
| | NHL | 60 | 299,441 | 406 | 62.65 | 1.42 | 60.60 | 61.45 | 62.70 | 63.91 | 64.53 | 299,441 | 436 | 61.04 | 3.78 | 56.71 | 59.40 | 62.14 | 63.73 | 64.45 |
| | NHL | 65 | 292,150 | 584 | 67.70 | 1.48 | 65.59 | 66.43 | 67.80 | 69.05 | 69.67 | 292,150 | 583 | 66.19 | 3.91 | 61.36 | 65.08 | 67.28 | 68.84 | 69.63 |
| | MM | 40 | 397,561 | 24 | 43.10 | 1.44 | 41.07 | 42.01 | 43.10 | 44.56 | 44.65 | 397,561 | 27 | 42.60 | 1.98 | 39.70 | 41.63 | 43.01 | 44.44 | 44.64 |
| | MM | 45 | 411,925 | 57 | 47.93 | 1.28 | 46.17 | 46.89 | 48.13 | 48.96 | 49.41 | 411,931 | 61 | 47.30 | 2.33 | 44.55 | 46.50 | 47.92 | 48.88 | 49.40 |
| | MM | 50 | 382,840 | 106 | 52.91 | 1.34 | 50.92 | 51.77 | 53.03 | 54.12 | 54.63 | 382,840 | 100 | 52.62 | 2.52 | 50.71 | 51.80 | 53.07 | 54.16 | 54.64 |
| | MM | 55 | 332,946 | 133 | 57.52 | 1.34 | 55.62 | 56.53 | 57.59 | 58.56 | 59.34 | 332,946 | 138 | 57.18 | 1.79 | 54.70 | 56.01 | 57.42 | 58.47 | 59.30 |
| | MM | 60 | 299,441 | 163 | 62.76 | 1.40 | 60.70 | 61.74 | 62.75 | 63.88 | 64.66 | 299,441 | 168 | 61.84 | 2.87 | 57.54 | 60.54 | 62.52 | 63.87 | 64.62 |
| | MM | 65 | 292,150 | 278 | 67.73 | 1.36 | 65.77 | 66.55 | 67.82 | 68.87 | 69.57 | 292,150 | 302 | 66.73 | 3.13 | 63.76 | 65.99 | 67.48 | 68.72 | 69.54 |
| | AML | 40 | 397,561 | 14 | 42.84 | 1.23 | 40.94 | 42.54 | 43.15 | 43.62 | 44.16 | 397,565 | 16 | 41.92 | 2.10 | 38.63 | 40.01 | 42.93 | 43.53 | 44.10 |
| | AML | 45 | 411,925 | 24 | 48.06 | 1.07 | 46.40 | 47.38 | 48.14 | 48.90 | 49.21 | 411,925 | 22 | 47.13 | 2.46 | 43.31 | 46.83 | 47.93 | 48.87 | 49.15 |
| | AML | 50 | 382,840 | 32 | 52.28 | 1.47 | 50.29 | 51.20 | 51.88 | 53.39 | 54.48 | 382,840 | 34 | 51.24 | 3.16 | 48.31 | 50.17 | 51.74 | 53.30 | 54.44 |
| | AML | 55 | 332,946 | 40 | 57.84 | 1.57 | 55.39 | 56.73 | 58.08 | 59.36 | 59.71 | 332,946 | 45 | 56.36 | 3.89 | 51.76 | 55.16 | 57.50 | 59.04 | 59.68 |
| | AML | 60 | 299,441 | 66 | 62.62 | 1.55 | 60.61 | 61.12 | 62.94 | 64.00 | 64.65 | 299,441 | 70 | 61.22 | 4.48 | 58.09 | 60.79 | 62.28 | 63.97 | 64.62 |
| | AML | 65 | 292,150 | 99 | 67.76 | 1.48 | 65.65 | 66.42 | 67.93 | 69.00 | 69.75 | 292,151 | 104 | 66.36 | 4.76 | 63.43 | 65.78 | 67.69 | 68.90 | 69.74 |

| | Cancer | Age | Baseline | | | | | | | | | Population health informed screening | | | | | | | | |
|---|---|---|---|---|---|---|---|---|---|---|---|---|---|---|---|---|---|---|---|---|
| | | | N | Cancers | Age - mean | Age - s.d. | Age - q10 | Age - q25 | Age - median | Age - q75 | Age - q90 | N | Cancers | Age - mean | Age - s.d. | Age - q10 | Age - q25 | Age - median | Age - q75 | Age - q90 |
| UK Biobank | Oesophagus | 55 | 67,266 | 24 | 57.93 | 1.36 | 55.98 | 56.85 | 58.21 | 58.85 | 59.69 | 67,266 | 27 | 56.84 | 2.62 | 52.58 | 55.67 | 57.54 | 58.79 | 59.66 |
| | Oesophagus | 60 | 75,871 | 49 | 62.88 | 1.37 | 60.94 | 61.79 | 62.96 | 64.13 | 64.72 | 75,871 | 57 | 61.06 | 3.60 | 56.19 | 59.63 | 61.88 | 63.88 | 64.61 |
| | Oesophagus | 65 | 98,019 | 93 | 67.64 | 1.43 | 65.48 | 66.55 | 67.63 | 68.88 | 69.38 | 98,019 | 101 | 65.23 | 4.13 | 60.13 | 63.04 | 66.55 | 68.21 | 69.30 |
| | Stomach | 55 | 67,266 | 15 | 58.41 | 0.80 | 57.60 | 57.95 | 58.21 | 58.96 | 59.46 | 67,266 | 18 | 57.27 | 2.40 | 53.56 | 56.88 | 58.04 | 58.77 | 59.33 |
| | Stomach | 60 | 75,871 | 37 | 62.72 | 1.54 | 60.46 | 61.55 | 62.63 | 64.13 | 64.73 | 75,871 | 40 | 62.13 | 2.23 | 59.08 | 60.46 | 62.29 | 64.00 | 64.66 |
| | Stomach | 65 | 98,019 | 65 | 67.97 | 1.34 | 65.85 | 67.13 | 68.30 | 69.05 | 69.52 | 98,019 | 80 | 66.45 | 3.07 | 62.04 | 64.82 | 67.47 | 68.74 | 69.39 |
| | Colorectal | 55 | 67,266 | 181 | 57.57 | 1.40 | 55.62 | 56.37 | 57.62 | 58.71 | 59.46 | 67,266 | 190 | 57.29 | 1.76 | 55.03 | 56.12 | 57.54 | 58.71 | 59.38 |
| | Colorectal | 60 | 75,871 | 233 | 62.66 | 1.43 | 60.64 | 61.38 | 62.79 | 63.96 | 64.55 | 75,871 | 252 | 62.06 | 2.17 | 59.29 | 60.79 | 62.55 | 63.80 | 64.55 |
| | Colorectal | 65 | 98,019 | 430 | 67.58 | 1.37 | 65.72 | 66.46 | 67.59 | 68.72 | 69.47 | 98,019 | 462 | 66.86 | 2.17 | 63.81 | 65.63 | 67.21 | 68.55 | 69.38 |
| | Liver | 55 | 67,266 | 21 | 57.97 | 1.22 | 56.29 | 57.12 | 58.46 | 58.71 | 59.13 | 67,266 | 19 | 57.35 | 2.31 | 55.06 | 56.54 | 58.46 | 58.83 | 59.14 |
| | Liver | 60 | 75,871 | 32 | 63.09 | 1.37 | 61.38 | 62.33 | 63.51 | 64.13 | 64.76 | 75,871 | 38 | 61.04 | 3.52 | 56.20 | 58.58 | 62.46 | 63.86 | 64.46 |
| | Liver | 65 | 98,019 | 64 | 67.89 | 1.27 | 66.26 | 66.94 | 67.97 | 68.90 | 69.44 | 98,019 | 79 | 65.26 | 4.26 | 58.68 | 63.59 | 66.88 | 68.68 | 69.16 |
| | Pancreas | 55 | 67,266 | 27 | 57.80 | 1.33 | 55.71 | 57.08 | 58.04 | 58.71 | 59.58 | 67,266 | 30 | 57.09 | 2.17 | 53.63 | 56.04 | 57.50 | 58.58 | 59.55 |
| | Pancreas | 60 | 75,871 | 57 | 62.97 | 1.19 | 60.85 | 62.55 | 63.21 | 63.88 | 64.26 | 75,871 | 59 | 62.61 | 1.93 | 60.33 | 62.29 | 63.21 | 63.88 | 64.20 |
| | Pancreas | 65 | 98,019 | 119 | 67.73 | 1.35 | 65.78 | 66.80 | 67.88 | 68.80 | 69.47 | 98,019 | 118 | 67.19 | 2.23 | 63.88 | 66.02 | 67.63 | 68.80 | 69.47 |
| | Lung | 55 | 67,266 | 77 | 57.84 | 1.43 | 55.56 | 56.71 | 58.21 | 58.88 | 59.63 | 67,266 | 102 | 56.25 | 2.75 | 51.89 | 53.99 | 56.79 | 58.52 | 59.54 |
| | Lung | 60 | 75,871 | 191 | 62.72 | 1.44 | 60.63 | 61.46 | 63.04 | 63.88 | 64.55 | 75,871 | 245 | 60.73 | 3.52 | 55.07 | 58.79 | 61.71 | 63.55 | 64.38 |
| | Lung | 65 | 98,019 | 344 | 67.68 | 1.43 | 65.72 | 66.63 | 67.63 | 68.88 | 69.63 | 98,019 | 492 | 65.10 | 3.92 | 60.13 | 63.04 | 65.96 | 68.05 | 69.30 |
| | Breast | 55 | 37,670 | 334 | 57.63 | 1.43 | 55.62 | 56.46 | 57.71 | 58.88 | 59.52 | 37,670 | 366 | 56.56 | 2.59 | 52.50 | 55.04 | 57.04 | 58.79 | 59.46 |
| | Breast | 60 | 42,413 | 491 | 62.61 | 1.45 | 60.55 | 61.38 | 62.71 | 63.88 | 64.63 | 42,413 | 517 | 60.73 | 3.17 | 56.29 | 58.79 | 61.38 | 63.30 | 64.30 |
| | Breast | 65 | 54,118 | 686 | 67.51 | 1.35 | 65.59 | 66.46 | 67.55 | 68.55 | 69.38 | 54,118 | 744 | 63.56 | 4.07 | 58.06 | 60.77 | 64.04 | 66.96 | 68.30 |
| | Corpus Uteri | 55 | 37,670 | 56 | 57.51 | 1.44 | 55.63 | 56.12 | 57.46 | 58.81 | 59.42 | 37,670 | 65 | 56.63 | 2.19 | 53.54 | 55.21 | 56.62 | 58.54 | 59.34 |
| | Corpus Uteri | 60 | 42,413 | 83 | 62.62 | 1.47 | 60.71 | 61.25 | 62.55 | 63.84 | 64.70 | 42,413 | 98 | 60.88 | 2.89 | 56.18 | 58.85 | 61.17 | 63.17 | 64.55 |
| | Corpus Uteri | 65 | 54,118 | 110 | 67.54 | 1.29 | 65.71 | 66.63 | 67.63 | 68.47 | 69.47 | 54,118 | 144 | 64.77 | 3.88 | 58.69 | 62.52 | 65.67 | 67.88 | 68.83 |
| | Ovary | 55 | 37,670 | 36 | 58.02 | 1.41 | 55.71 | 56.91 | 58.42 | 59.23 | 59.63 | 37,670 | 31 | 57.18 | 2.33 | 53.54 | 55.71 | 58.12 | 59.21 | 59.46 |
| | Ovary | 60 | 42,413 | 59 | 62.82 | 1.43 | 60.94 | 61.67 | 62.71 | 64.21 | 64.73 | 42,413 | 52 | 62.15 | 2.28 | 59.06 | 60.75 | 62.59 | 64.09 | 64.79 |
| | Ovary | 65 | 54,118 | 82 | 67.40 | 1.39 | 65.39 | 66.11 | 67.47 | 68.30 | 69.37 | 54,118 | 87 | 66.16 | 2.57 | 62.85 | 64.92 | 66.38 | 68.00 | 69.28 |
| | Prostate | 55 | 29,596 | 212 | 57.75 | 1.47 | 55.46 | 56.54 | 57.88 | 59.04 | 59.63 | 29,596 | 222 | 57.50 | 1.77 | 54.87 | 56.37 | 57.75 | 59.04 | 59.63 |
| | Prostate | 60 | 33,458 | 381 | 62.82 | 1.39 | 60.71 | 61.71 | 62.88 | 64.04 | 64.72 | 33,458 | 397 | 62.32 | 2.10 | 59.38 | 61.21 | 62.71 | 63.96 | 64.66 |
| | Prostate | 65 | 43,901 | 840 | 67.62 | 1.38 | 65.72 | 66.55 | 67.88 | 68.72 | 69.38 | 43,901 | 829 | 66.83 | 2.43 | 63.45 | 65.55 | 67.47 | 68.63 | 69.38 |
| | Kidney | 55 | 67,266 | 31 | 57.71 | 1.56 | 55.54 | 56.46 | 57.79 | 59.08 | 59.79 | 67,266 | 35 | 57.06 | 2.23 | 54.29 | 55.50 | 57.46 | 58.96 | 59.79 |
| | Kidney | 60 | 75,871 | 56 | 63.11 | 1.38 | 60.92 | 61.94 | 63.46 | 64.21 | 64.80 | 75,871 | 64 | 61.68 | 3.22 | 56.33 | 59.88 | 63.34 | 63.90 | 64.69 |
| | Kidney | 65 | 98,019 | 127 | 67.76 | 1.29 | 65.82 | 66.96 | 67.88 | 68.76 | 69.47 | 98,019 | 118 | 66.69 | 2.85 | 63.38 | 65.36 | 67.55 | 68.70 | 69.41 |
| | Bladder | 55 | 67,266 | 60 | 57.87 | 1.48 | 55.53 | 56.93 | 58.12 | 59.15 | 59.71 | 67,266 | 61 | 57.60 | 1.79 | 55.21 | 56.62 | 57.88 | 59.13 | 59.71 |
| | Bladder | 60 | 75,871 | 85 | 62.75 | 1.41 | 60.66 | 61.71 | 62.71 | 64.04 | 64.68 | 75,871 | 95 | 62.12 | 2.12 | 59.58 | 60.75 | 62.55 | 63.84 | 64.60 |
| | Bladder | 65 | 98,019 | 195 | 67.68 | 1.34 | 65.80 | 66.63 | 67.80 | 68.76 | 69.47 | 98,019 | 207 | 67.13 | 2.02 | 64.35 | 65.96 | 67.47 | 68.72 | 69.47 |
| | Brain | 55 | 67,266 | 29 | 57.59 | 1.29 | 55.86 | 56.46 | 57.62 | 58.46 | 59.39 | 67,266 | 27 | 57.41 | 1.57 | 55.24 | 56.29 | 57.62 | 58.58 | 59.49 |
| | Brain | 60 | 75,871 | 43 | 62.70 | 1.45 | 60.88 | 61.55 | 62.46 | 64.09 | 64.72 | 75,871 | 43 | 62.39 | 2.00 | 60.14 | 61.42 | 62.29 | 64.09 | 64.72 |
| | Brain | 65 | 98,019 | 64 | 67.48 | 1.28 | 65.57 | 66.53 | 67.51 | 68.40 | 69.19 | 98,019 | 60 | 66.80 | 2.04 | 64.51 | 65.25 | 67.26 | 68.40 | 69.22 |
| | NHL | 55 | 67,266 | 52 | 58.03 | 1.45 | 55.88 | 56.77 | 58.25 | 59.29 | 59.79 | 67,266 | 57 | 57.16 | 2.63 | 52.87 | 56.04 | 58.12 | 59.21 | 59.79 |
| | NHL | 60 | 75,871 | 91 | 62.93 | 1.40 | 60.71 | 61.88 | 63.21 | 64.00 | 64.63 | 75,871 | 94 | 61.69 | 2.97 | 58.26 | 60.15 | 62.50 | 63.88 | 64.63 |
| | NHL | 65 | 98,019 | 162 | 67.63 | 1.41 | 65.72 | 66.55 | 67.55 | 68.94 | 69.63 | 98,019 | 171 | 66.42 | 3.28 | 62.46 | 65.25 | 67.30 | 68.68 | 69.63 |
| | MM | 55 | 67,266 | 29 | 57.37 | 1.34 | 55.46 | 56.29 | 57.54 | 58.37 | 59.39 | 67,266 | 28 | 57.40 | 1.38 | 55.36 | 56.42 | 57.58 | 58.39 | 59.40 |
| | MM | 60 | 75,871 | 29 | 62.43 | 1.45 | 60.92 | 61.21 | 62.38 | 63.30 | 64.66 | 75,871 | 29 | 62.17 | 1.84 | 59.46 | 61.04 | 62.38 | 63.30 | 64.66 |
| | MM | 65 | 98,019 | 98 | 67.61 | 1.40 | 65.52 | 66.57 | 67.72 | 68.70 | 69.49 | 98,019 | 92 | 67.29 | 2.14 | 65.04 | 66.53 | 67.72 | 68.70 | 69.54 |
| | AML | 55 | 67,266 | 7 | 57.26 | 1.02 | 55.76 | 56.50 | 57.54 | 58.04 | 58.31 | 67,266 | 8 | 56.96 | 1.24 | 55.46 | 55.77 | 57.37 | 57.96 | 58.28 |
| | AML | 60 | 75,871 | 13 | 62.68 | 1.31 | 60.89 | 62.29 | 62.71 | 63.55 | 64.26 | 75,871 | 15 | 62.03 | 2.06 | 59.09 | 60.92 | 62.71 | 63.42 | 64.15 |
| | AML | 65 | 98,019 | 10 | 68.22 | 1.18 | 66.41 | 67.74 | 68.47 | 68.92 | 69.64 | 98,019 | 17 | 65.23 | 3.84 | 59.96 | 62.96 | 65.96 | 68.47 | 69.28 |

**ST7:** Baseline risk comparison to Age-sex model on the UK Biobank

| Cancer | Spearman Corr. | Pvalue | AUC Breslow | AUC UKB-AgeSex |
|---|---|---|---|---|
| Oesophagus | 0.964 | 0.000 | 0.715 | 0.722 |
| Stomach | 0.986 | 0.000 | 0.682 | 0.687 |
| Colorectal | 0.991 | 0.000 | 0.639 | 0.640 |
| Liver | 0.977 | 0.000 | 0.701 | 0.701 |
| Pancreas | 0.991 | 0.000 | 0.657 | 0.660 |
| Lung | 0.968 | 0.000 | 0.668 | 0.673 |
| Melanoma | 0.836 | 0.000 | 0.577 | 0.591 |
| Breast | 0.615 | 0.000 | 0.535 | 0.547 |
| Cervix Uteri | -0.777 | 0.000 | 0.472 | 0.513 |
| Corpus Uteri | 0.930 | 0.000 | 0.556 | 0.554 |
| Ovary | 0.995 | 0.000 | 0.575 | 0.576 |
| Prostate | 1.000 | 0.000 | 0.651 | 0.651 |
| Testis | -0.494 | 0.000 | 0.528 | 0.509 |
| Kidney | 0.938 | 0.000 | 0.668 | 0.675 |
| Bladder | 0.998 | 0.000 | 0.751 | 0.751 |
| Brain | 0.969 | 0.000 | 0.588 | 0.594 |
| Thyroid | 0.734 | 0.000 | 0.620 | 0.663 |
| Non-Hodgkin Lymphoma | 0.959 | 0.000 | 0.629 | 0.634 |
| Multiple Myeloma | 0.981 | 0.000 | 0.659 | 0.662 |
| AML | 0.933 | 0.000 | 0.624 | 0.639 |
| Other | 0.993 | 0.000 | 0.652 | 0.654 |
| Death | 0.926 | 0.000 | 0.695 | 0.709 |

**Notes:** Corr := Correlation, Breslow := Baseline hazard estimate based on Danish data, UKB-AgeSex := Age-Sex model fitted on UKB data

**ST8:** Proportional hazards assumption for the overall predictor.

| | Cancer | PH-Assumption Test | |
|---|---|---|---|
| | | Chi2 Statistic | P-value |
| Denmark | Oesophagus | 13.091 | 0.000 |
| | Stomach | 10.060 | 0.002 |
| | Colorectal | 8.734 | 0.003 |
| | Liver | 0.833 | 0.361 |
| | Pancreas | 4.667 | 0.031 |
| | Lung | 0.317 | 0.573 |
| | Melanoma | 0.087 | 0.769 |
| | Breast | 1.433 | 0.231 |
| | Cervix Uteri | 0.319 | 0.572 |
| | Corpus Uteri | 2.875 | 0.090 |
| | Ovary | 0.875 | 0.350 |
| | Prostate | 121.108 | 0.000 |
| | Testis | 1.310 | 0.252 |
| | Kidney | 12.333 | 0.000 |
| | Bladder | 2.121 | 0.145 |
| | Brain | 6.961 | 0.008 |
| | Thyroid | 0.040 | 0.841 |
| | Non-Hodgkin Lyr | 5.575 | 0.018 |
| | Multiple Myeloma | 10.409 | 0.001 |
| | AML | 0.008 | 0.928 |
| | Other | 4.097 | 0.043 |
| | Death | 381.505 | 0.000 |
| UK Biobank | Oesophagus | 2.015 | 0.156 |
| | Stomach | 1.752 | 0.186 |
| | Colorectal | 0.436 | 0.509 |
| | Liver | 4.105 | 0.043 |
| | Pancreas | 0.007 | 0.931 |
| | Lung | 3.013 | 0.083 |
| | Melanoma | 0.056 | 0.813 |
| | Breast | 0.100 | 0.752 |
| | Cervix Uteri | 0.004 | 0.949 |
| | Corpus Uteri | 2.324 | 0.127 |
| | Ovary | 2.209 | 0.137 |
| | Prostate | 5.032 | 0.025 |
| | Testis | 0.494 | 0.482 |
| | Kidney | 3.881 | 0.049 |
| | Bladder | 0.012 | 0.912 |
| | Brain | 1.565 | 0.211 |
| | Thyroid | 1.297 | 0.255 |
| | Non-Hodgkin Lyr | 0.237 | 0.626 |
| | Multiple Myeloma | 0.697 | 0.404 |
| | AML | 2.888 | 0.089 |
| | Other | 0.985 | 0.321 |
| | Death | 67.945 | 0.000 |

**Notes:** PH := Proportional Hazards

#### **SF1:** Age-sex adjusted concordance for the development data.

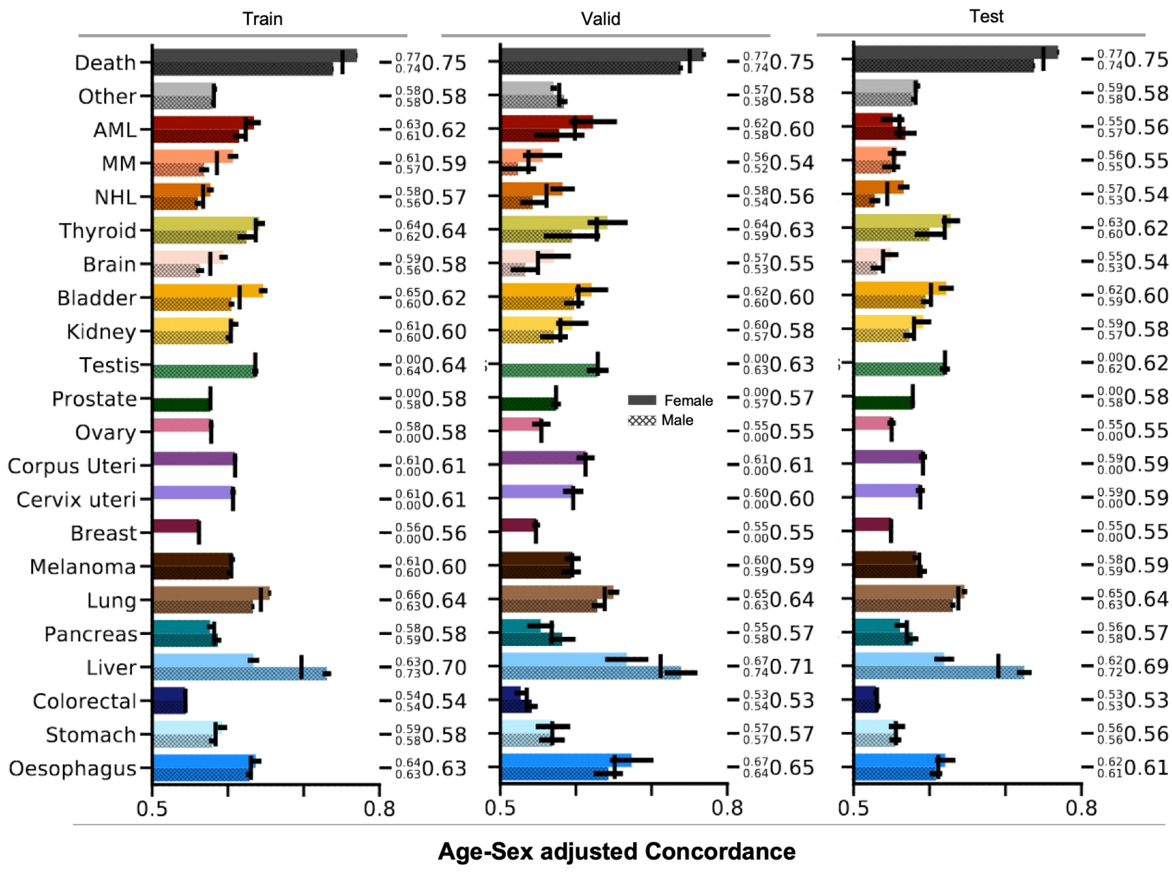

**supplementary figure 1:**

Concordance index for the development data on the Danish cohort.

#### **SFC-X:** Summary plots

a) Age-sex Incidence for the Danish and UKB cohort.
b) Model estimates for underlying baseline risk.
c) Forest plot based on HPD-90% – coloured for significant effects based on HPD-95%. A maximum of 30 associations are shown.
d) ROC-AUC curve and calibration plot for each decentile.
e) Kaplan-Meier curve for the top 1% risk quantile between our predictions and an Age-Sex baseline for the Danish and UKB cohort.
f) Lower/Upper quantiles of the 5-Year absolute risk as evaluated on the Danish and UKB cohort.

#### **SFC1:** Oesophagus

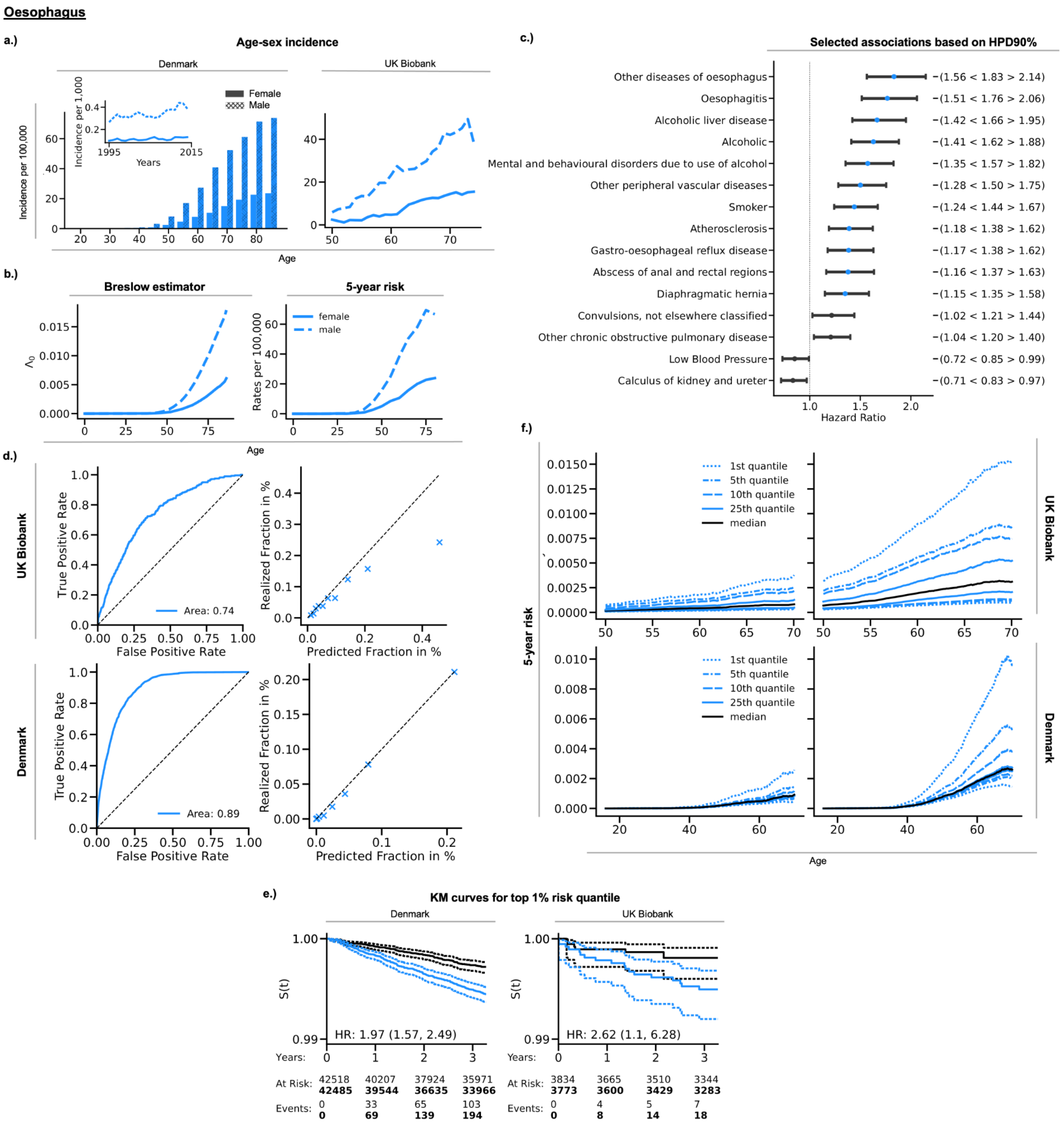

### **SFC2:** Stomach

**Stomach**

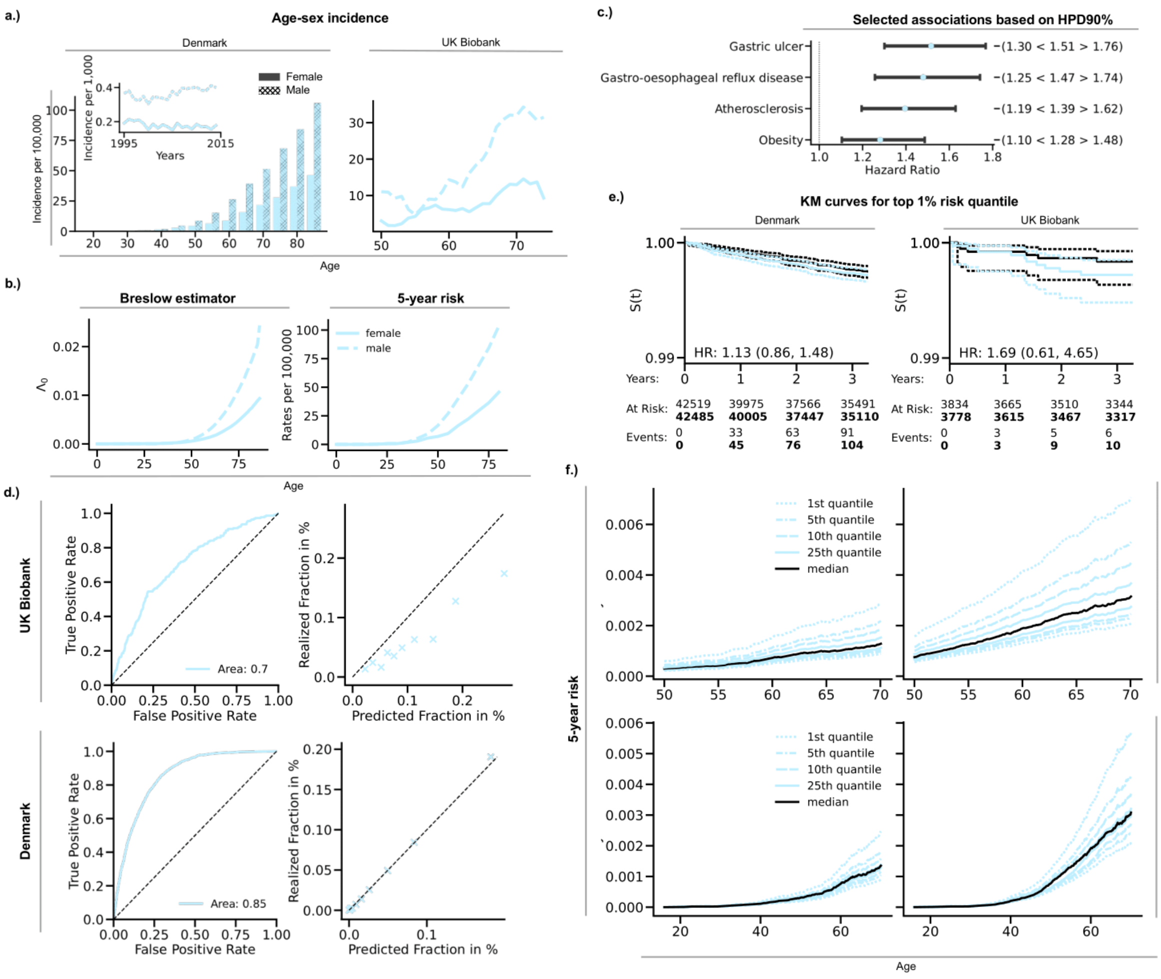

### **SFC3:** Colorectal

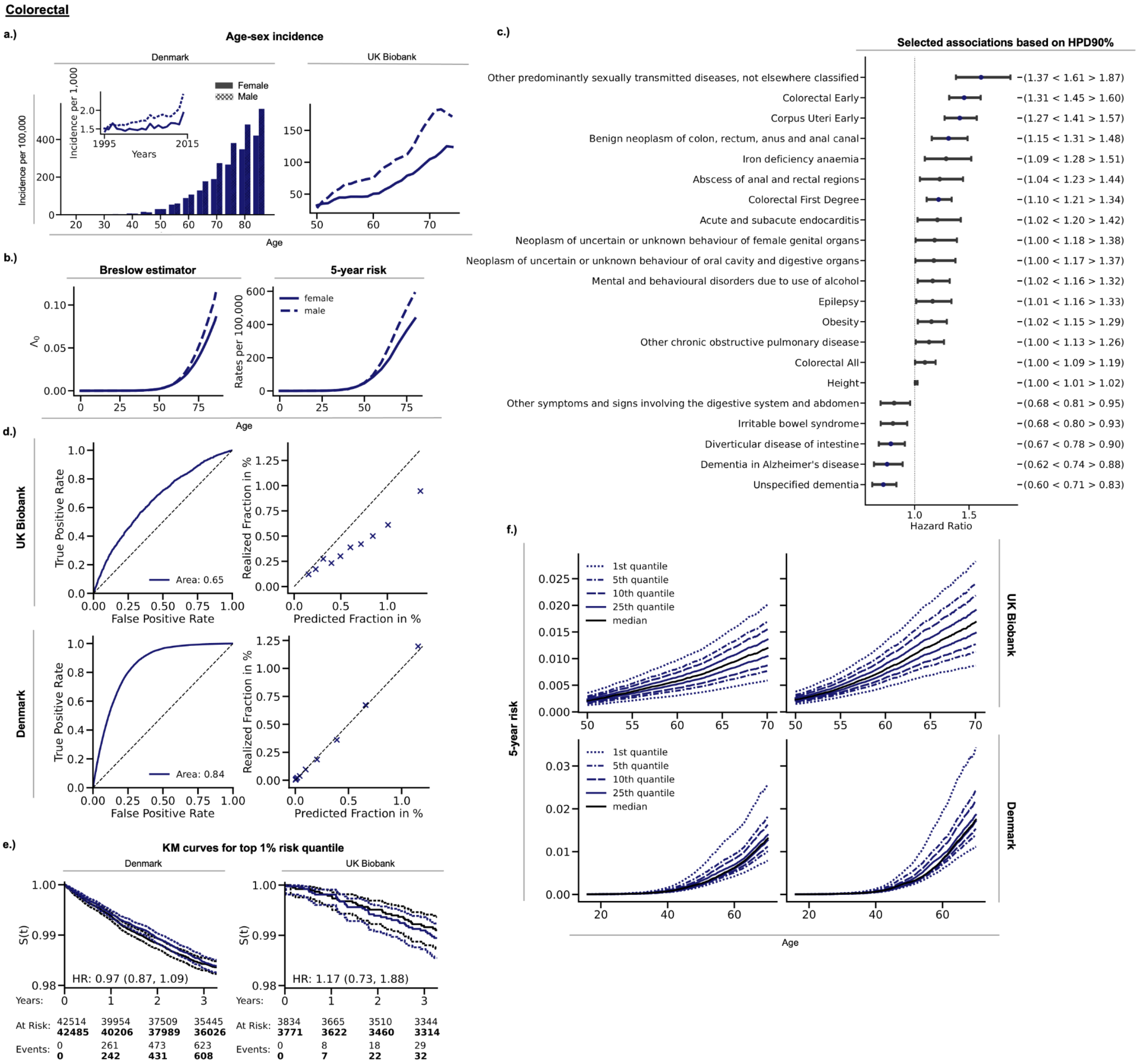

### **SFC4:** Liver

**Liver**

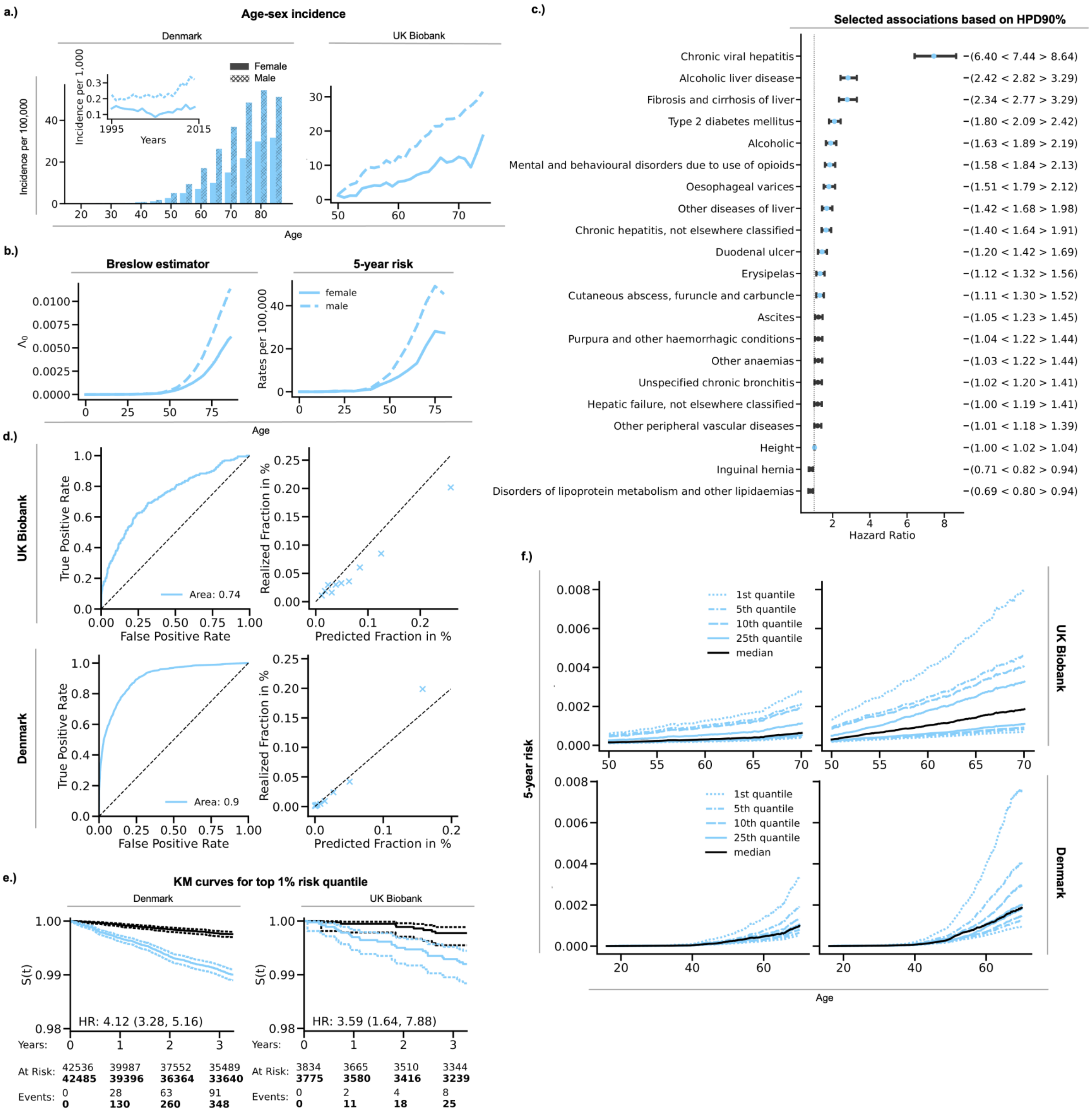

### **SFC5:** Pancreas

**Pancreas**

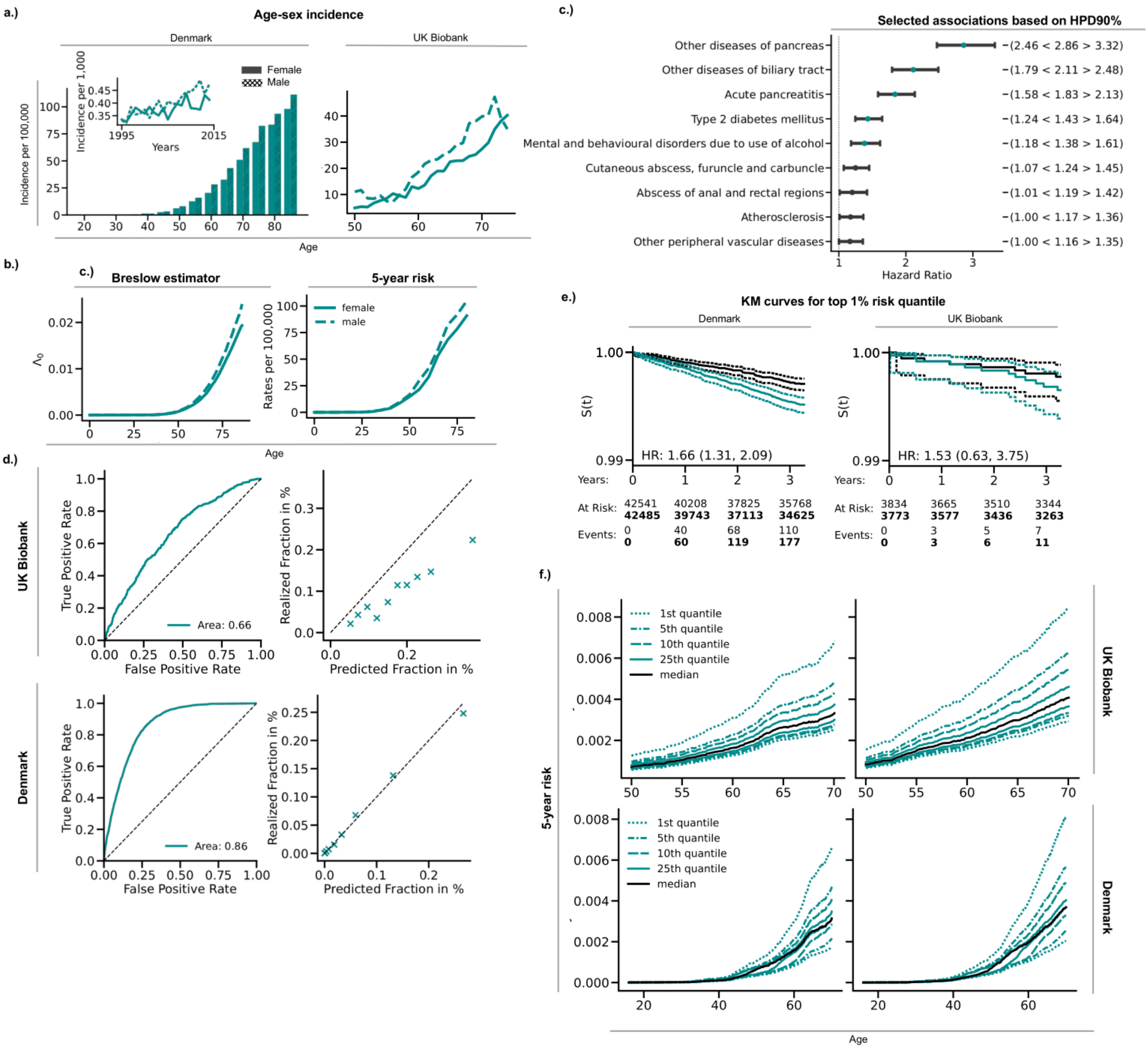

### **SFC6:** Lung

**Lung**

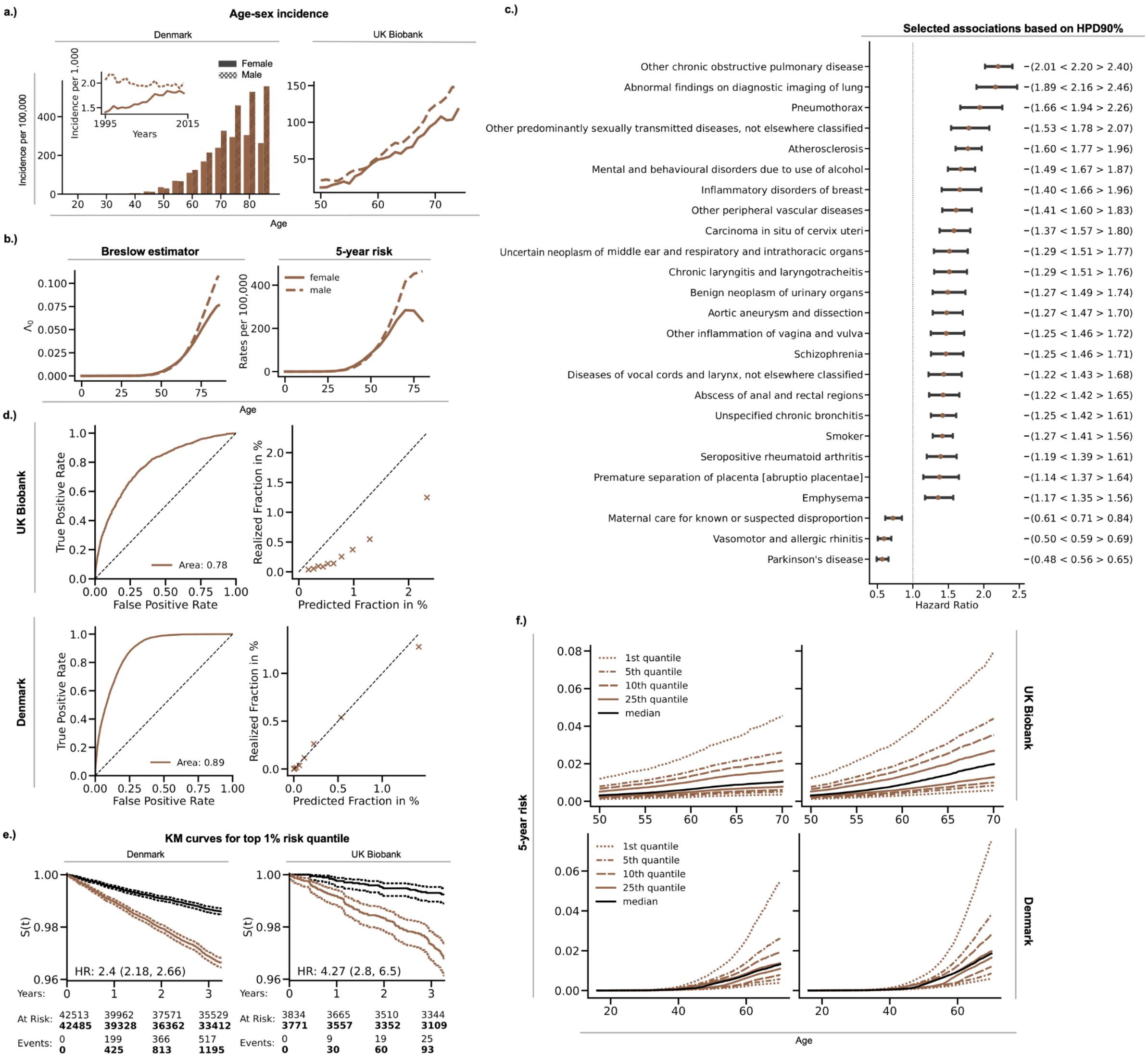

### **SFC7:** Melanoma

**Melanoma**

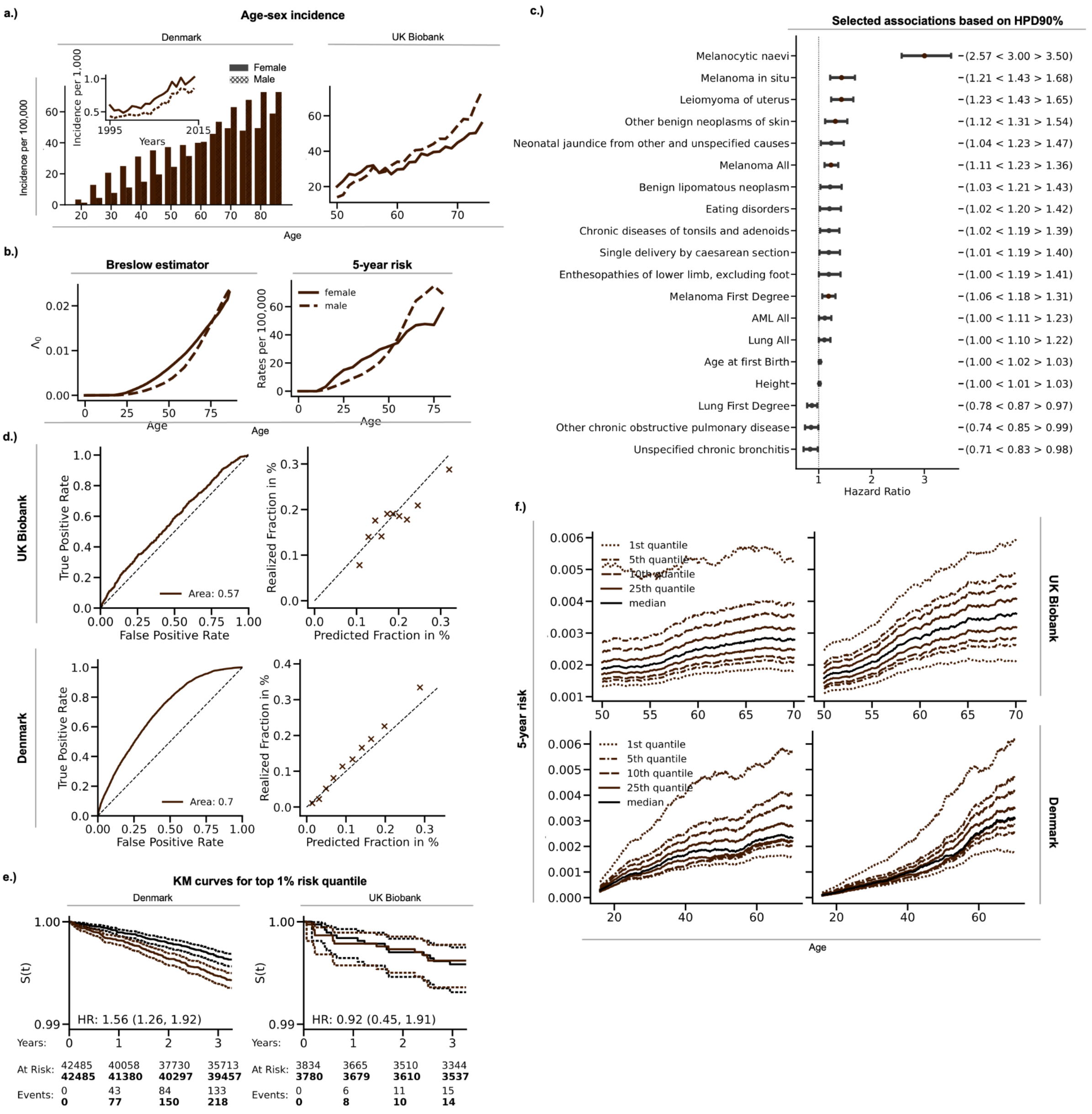

### **SFC8:** Breast

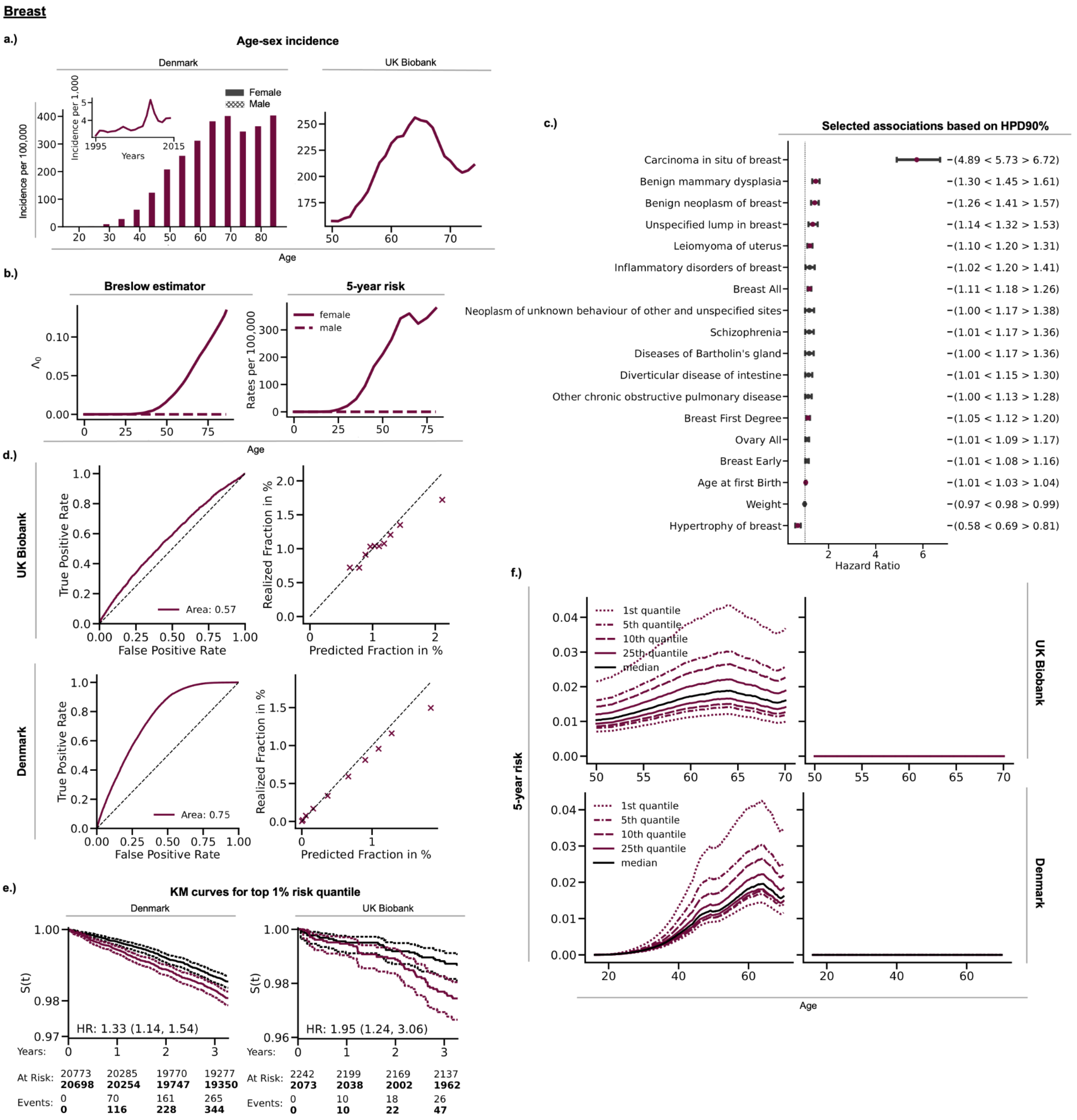

Breast
a.)
Age-sex incidence
Denmark
UK Biobank
Female
Male
Incidence per 100,000
Incidence per 1,000
1995
2015
Years
Age
b.)
Breslow estimator
5-year risk
female
male
Rates per 100,000
Age
c.)
Selected associations based on HPD90%
Carcinoma in situ of breast −(4.89 < 5.73 > 6.72)
Benign mammary dysplasia −(1.30 < 1.45 > 1.61)
Benign neoplasm of breast −(1.26 < 1.41 > 1.57)
Unspecified lump in breast −(1.14 < 1.32 > 1.53)
Leiomyoma of uterus −(1.10 < 1.20 > 1.31)
Inflammatory disorders of breast −(1.02 < 1.20 > 1.41)
Breast All −(1.11 < 1.18 > 1.26)
Neoplasm of unknown behaviour of other and unspecified sites −(1.00 < 1.17 > 1.38)
Schizophrenia −(1.01 < 1.17 > 1.36)
Diseases of Bartholin's gland −(1.00 < 1.17 > 1.36)
Diverticular disease of intestine −(1.01 < 1.15 > 1.30)
Other chronic obstructive pulmonary disease −(1.00 < 1.13 > 1.28)
Breast First Degree −(1.05 < 1.12 > 1.20)
Ovary All −(1.01 < 1.09 > 1.17)
Breast Early −(1.01 < 1.08 > 1.16)
Age at first Birth −(1.01 < 1.03 > 1.04)
Weight −(0.97 < 0.98 > 0.99)
Hypertrophy of breast −(0.58 < 0.69 > 0.81)
Hazard Ratio
d.)
UK Biobank
Denmark
True Positive Rate
False Positive Rate
Area: 0.57
Area: 0.75
Realized Fraction in %
Predicted Fraction in %
e.)
KM curves for top 1% risk quantile
Denmark
UK Biobank
S(t)
HR: 1.33 (1.14, 1.54)
HR: 1.95 (1.24, 3.06)
Years: 0 1 2 3
At Risk: 20773 20285 19770 19277
20698 20254 19747 19350
Events: 0 70 161 265
0 116 228 344
At Risk: 2242 2199 2169 2137
2073 2038 2002 1962
Events: 0 10 18 26
0 10 22 47
f.)
5-year risk
1st quantile
5th quantile
10th quantile
25th quantile
median
UK Biobank
Denmark
Age

### **SFC9:** Cervix Uteri

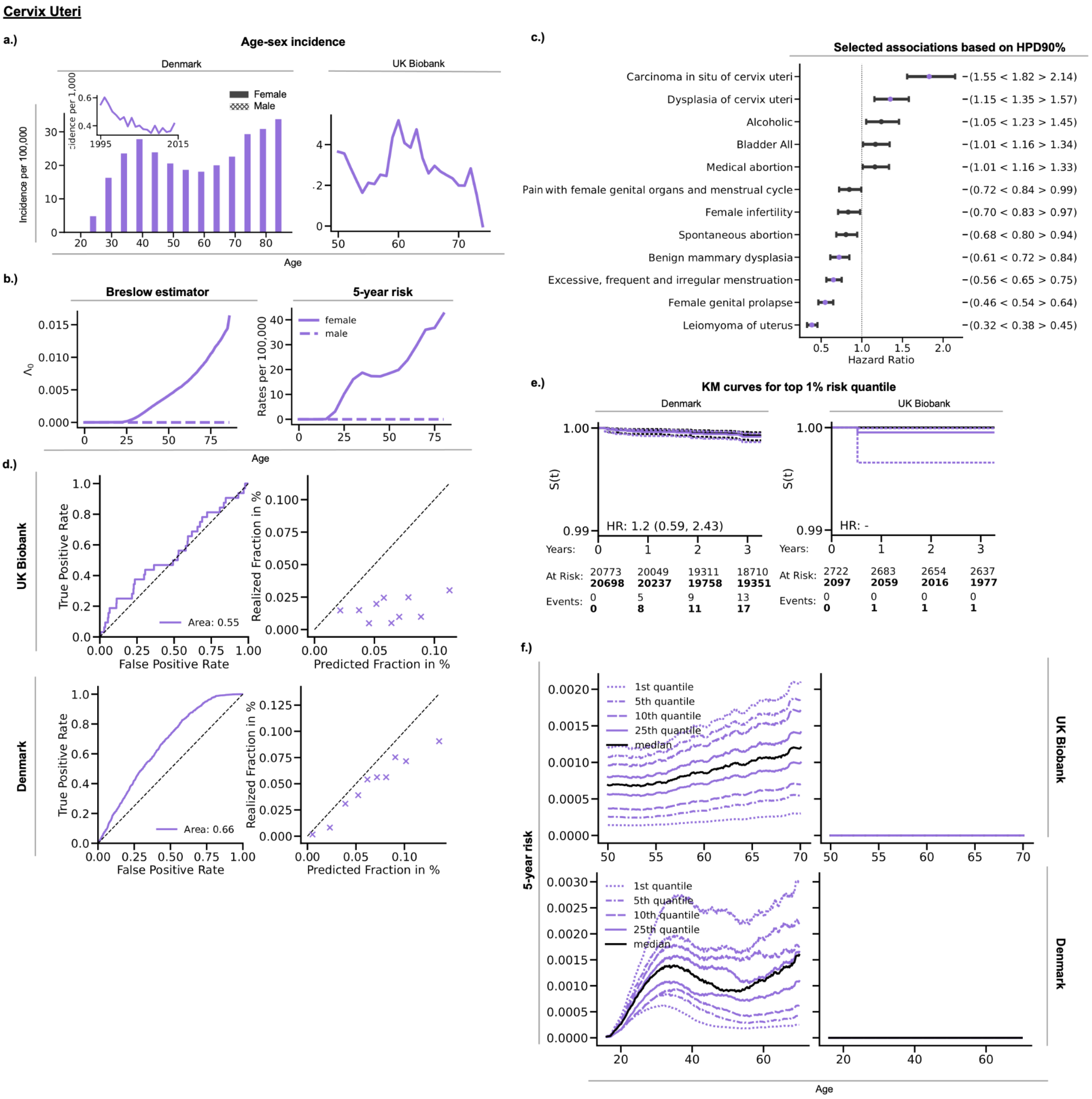

### **SFC10:** Corpus Uteri

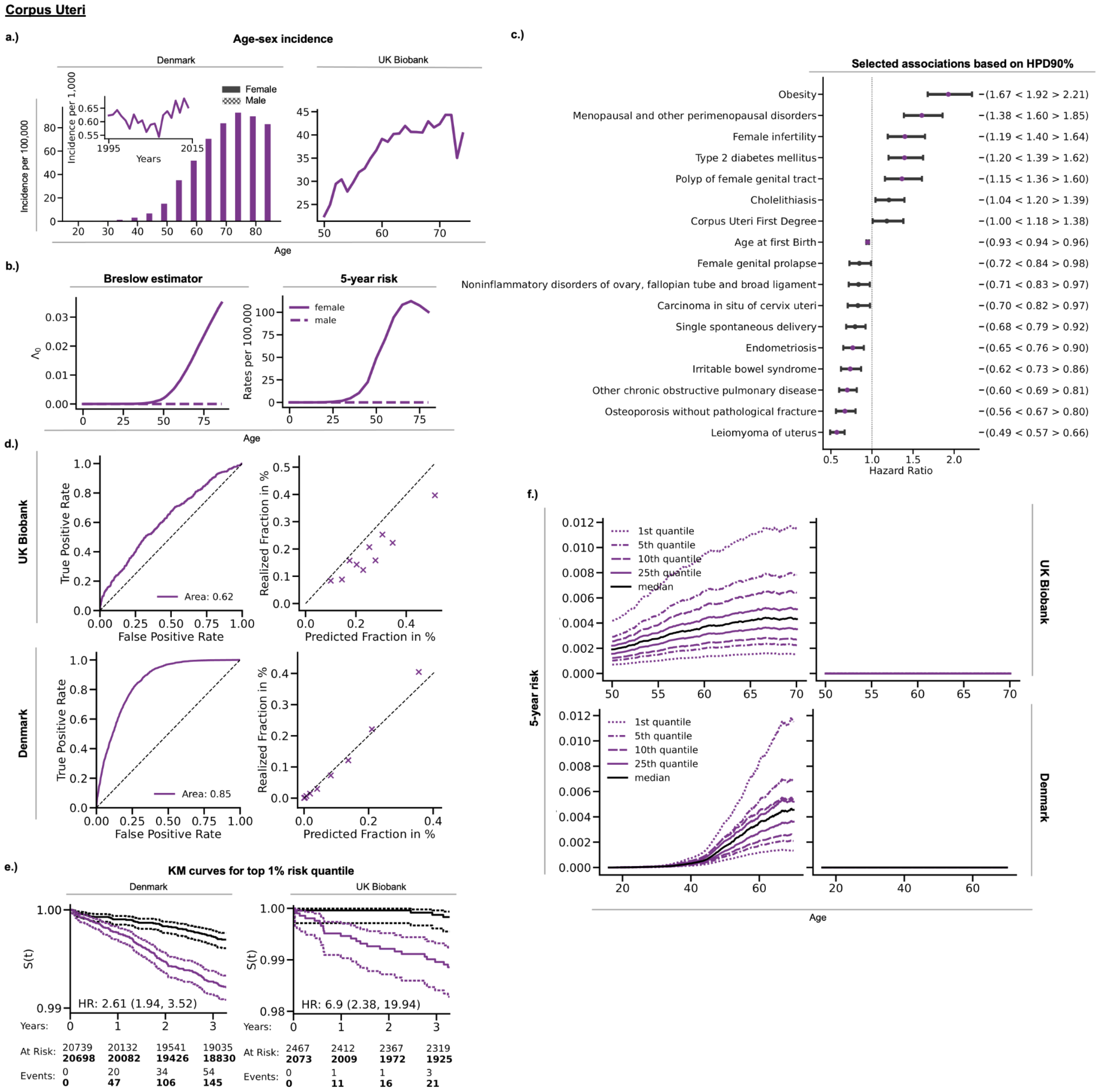

### **SFC11:** Ovary

**Ovary**

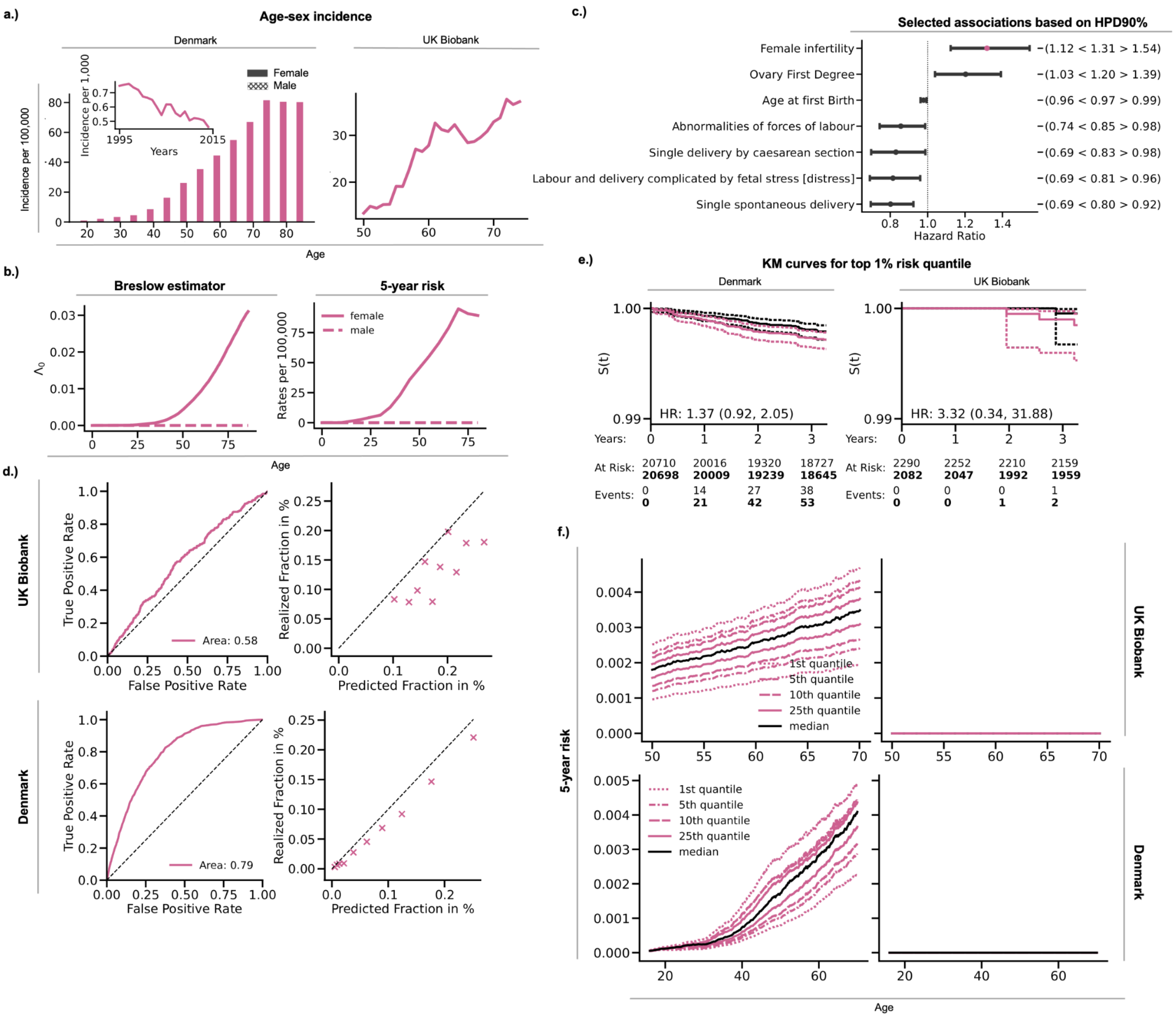

### **SFC12:** Prostate

**Prostate**

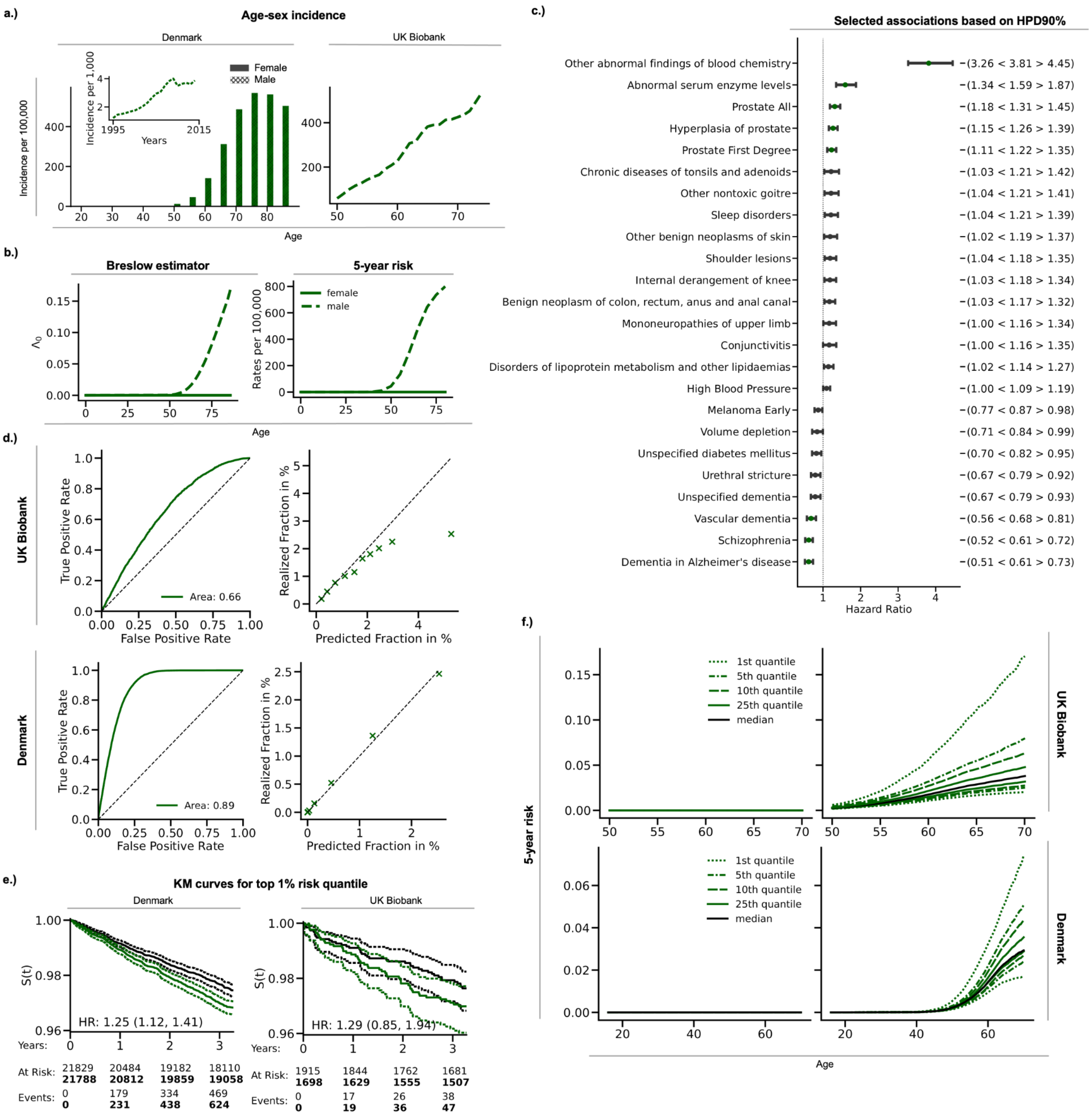

### **SFC13:** Testis

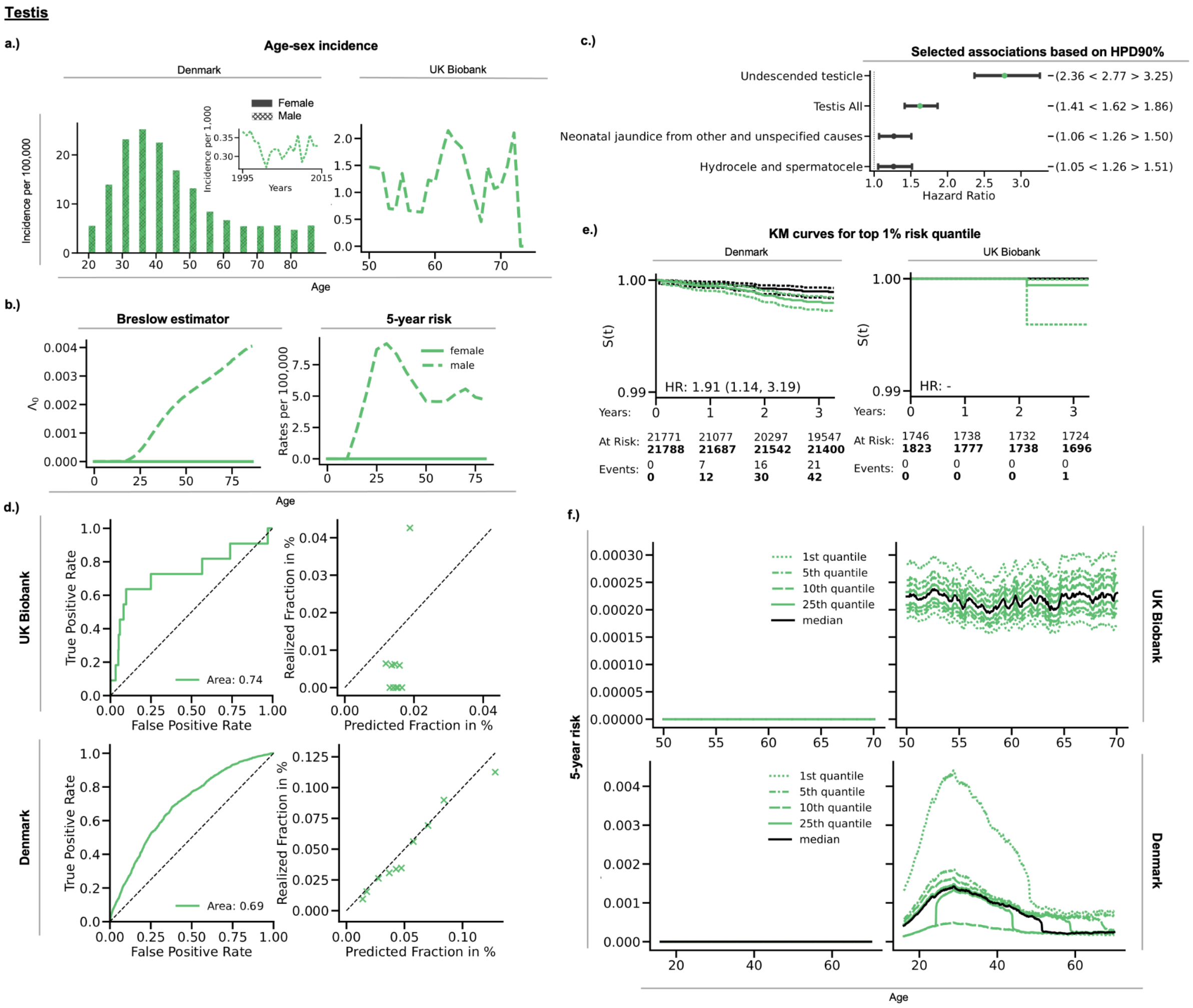

#### **SFC14:** Kidney

**Kidney**

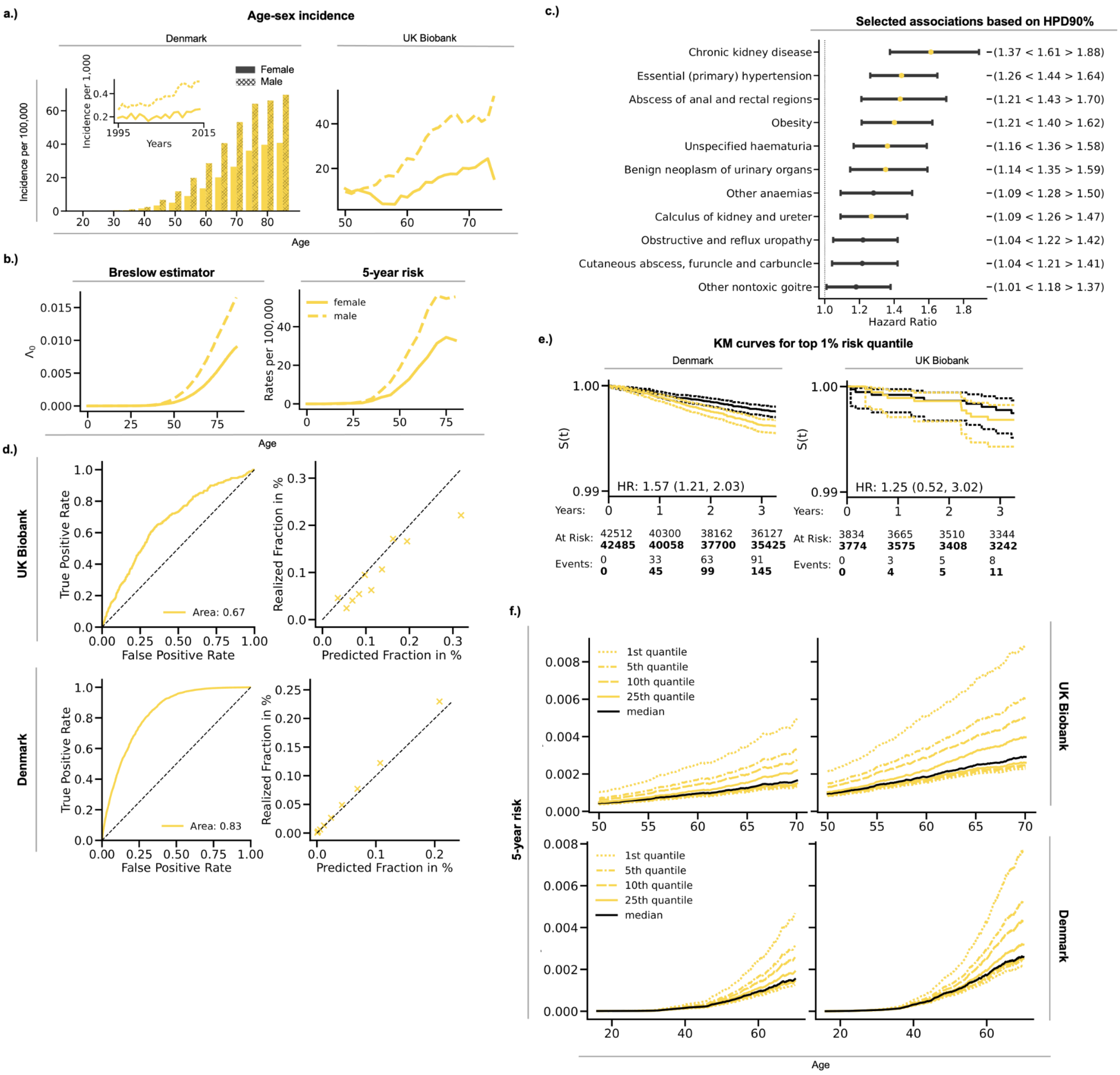

### **SFC15:** Bladder

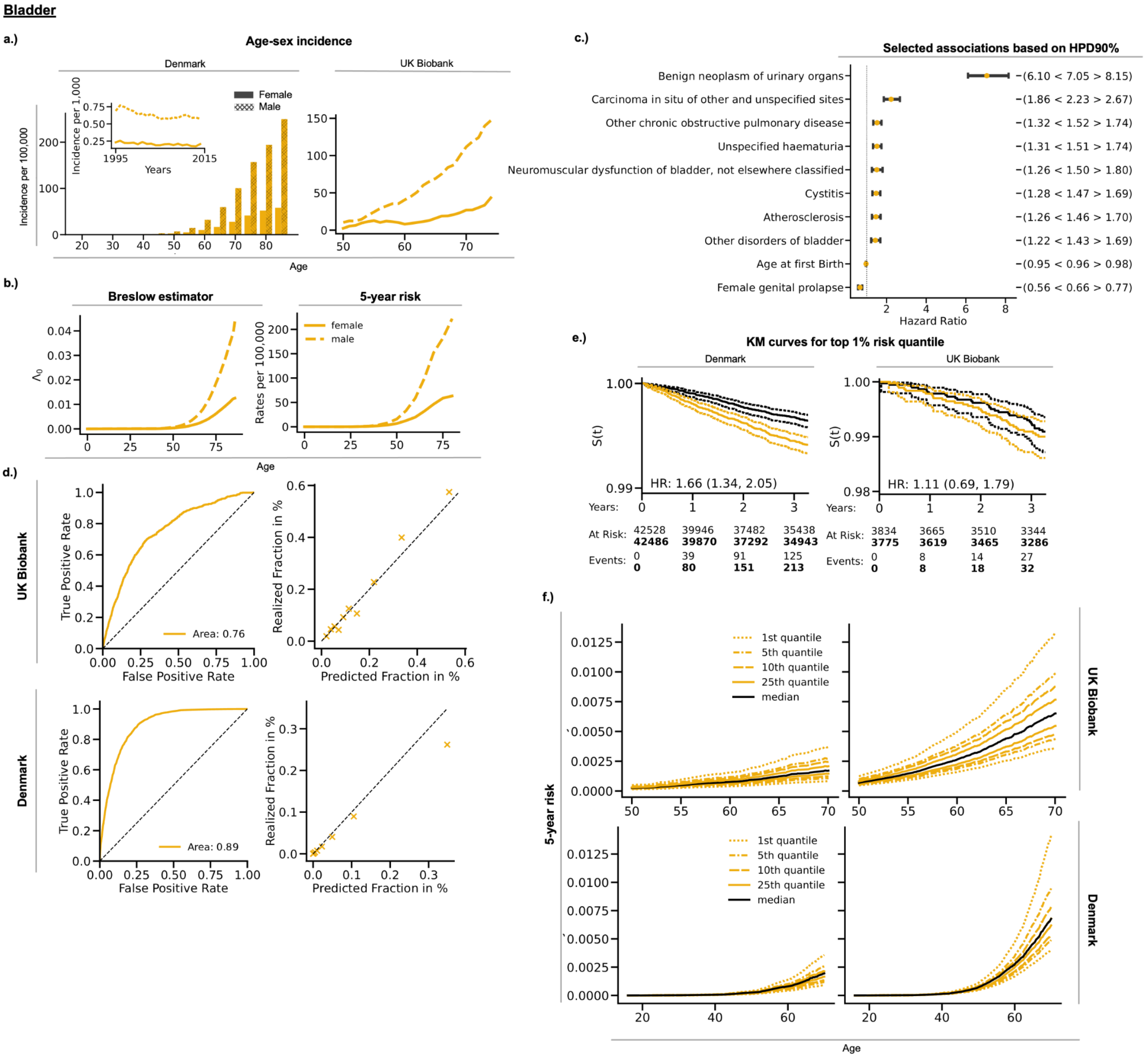

### **SFC16:** Brain

**Brain**

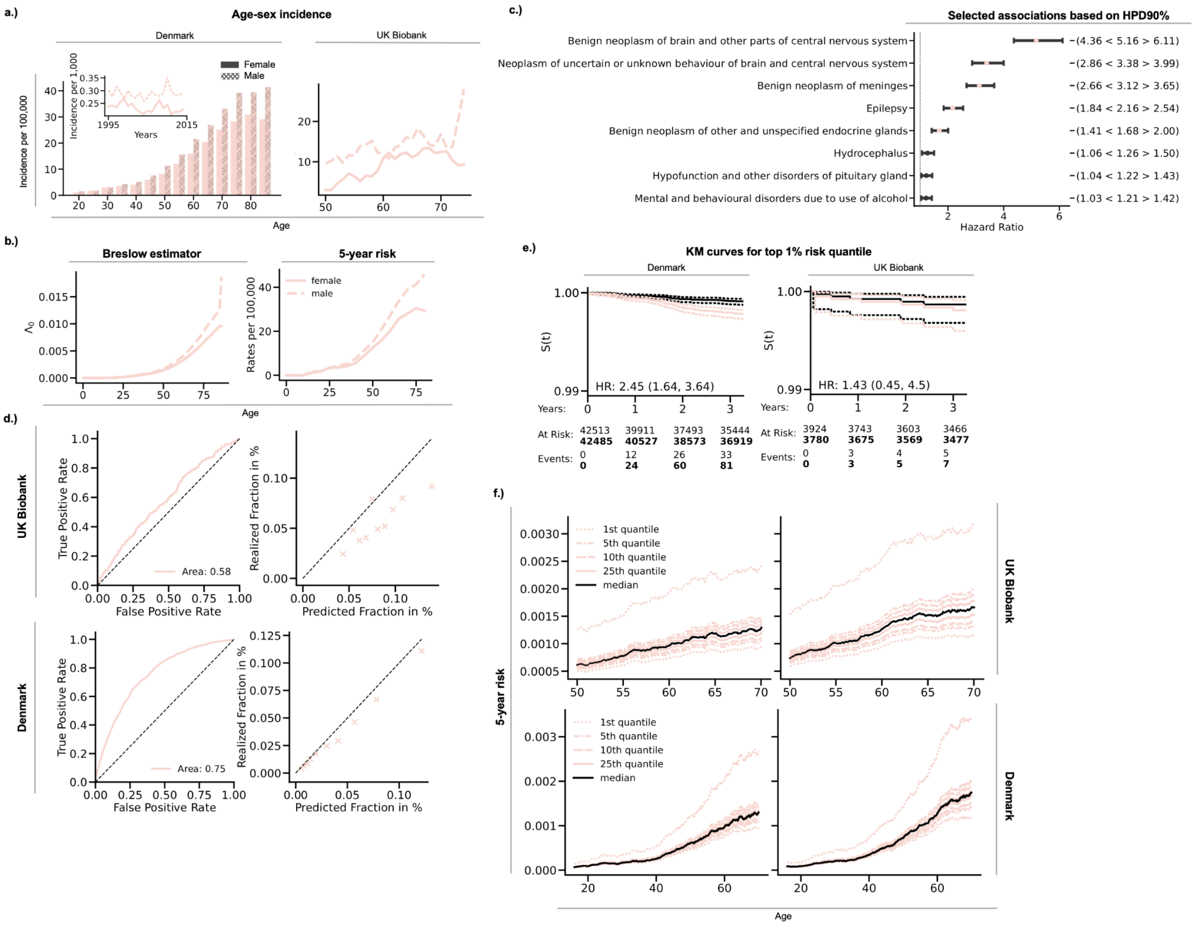

### **SFC17:** Thyroid

**Thyroid**

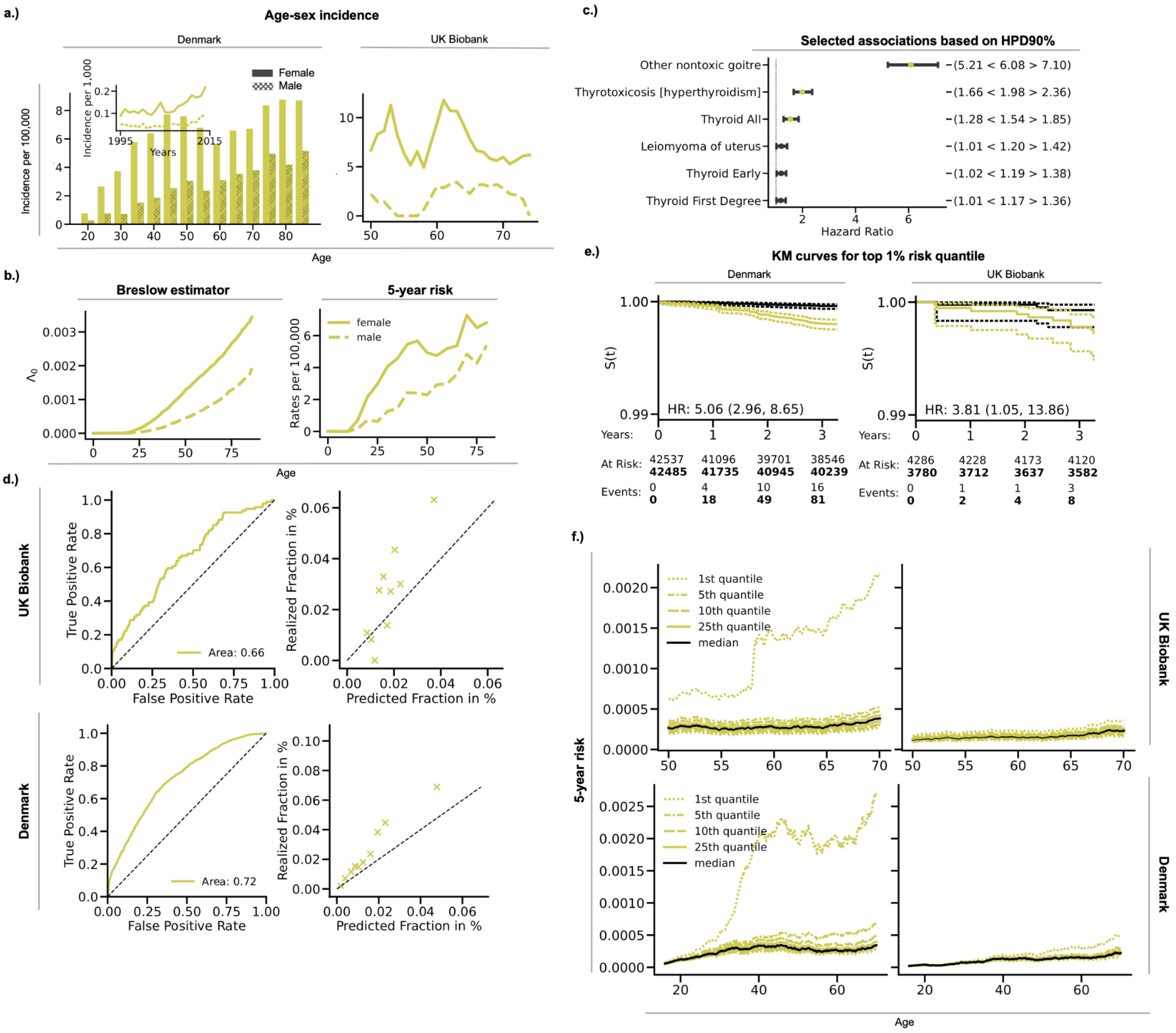

### **SFC18:** Non-hodgkin lymphoma

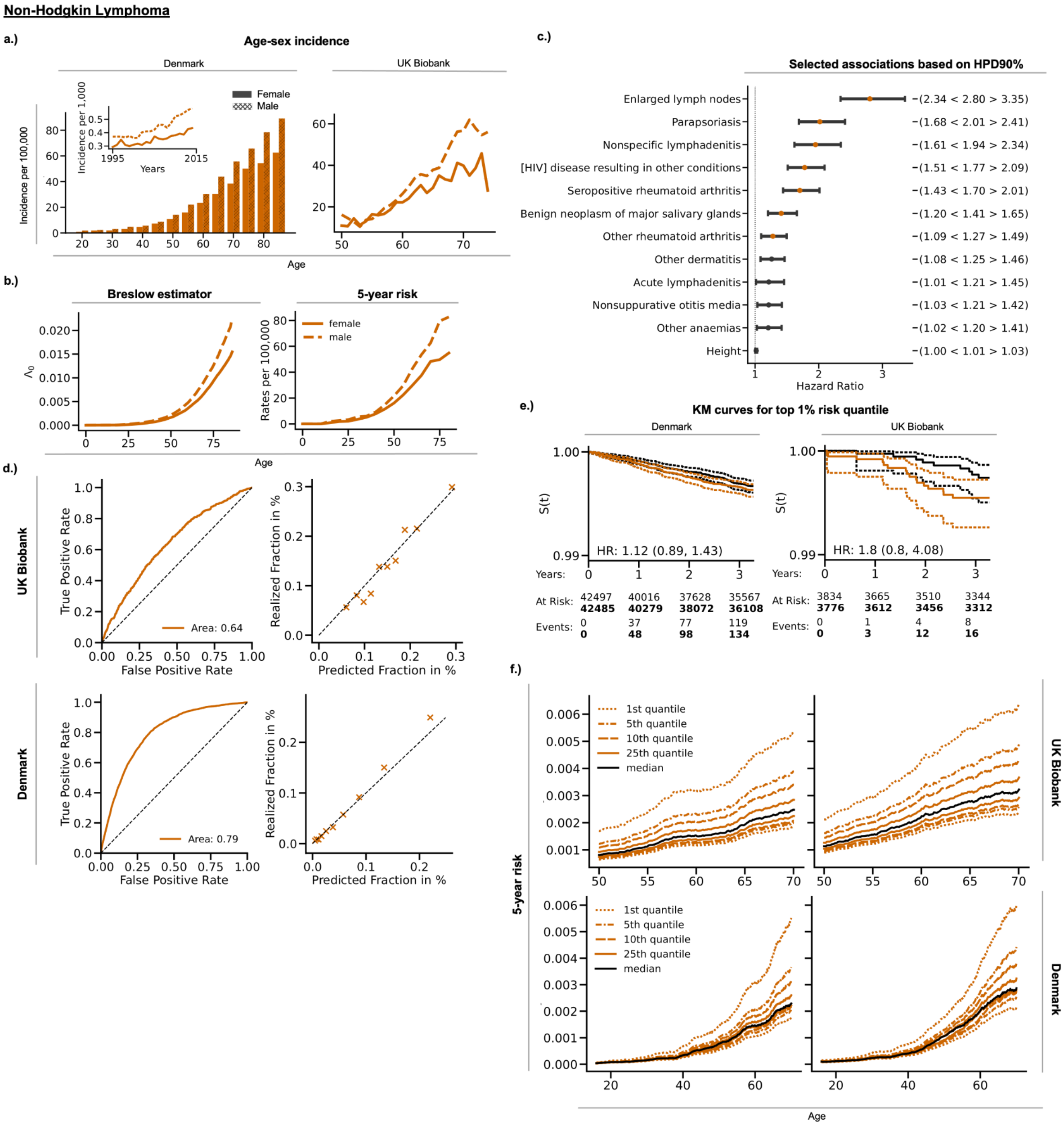

### **SFC19:** Multiple Myeloma

**Multiple Myeloma**

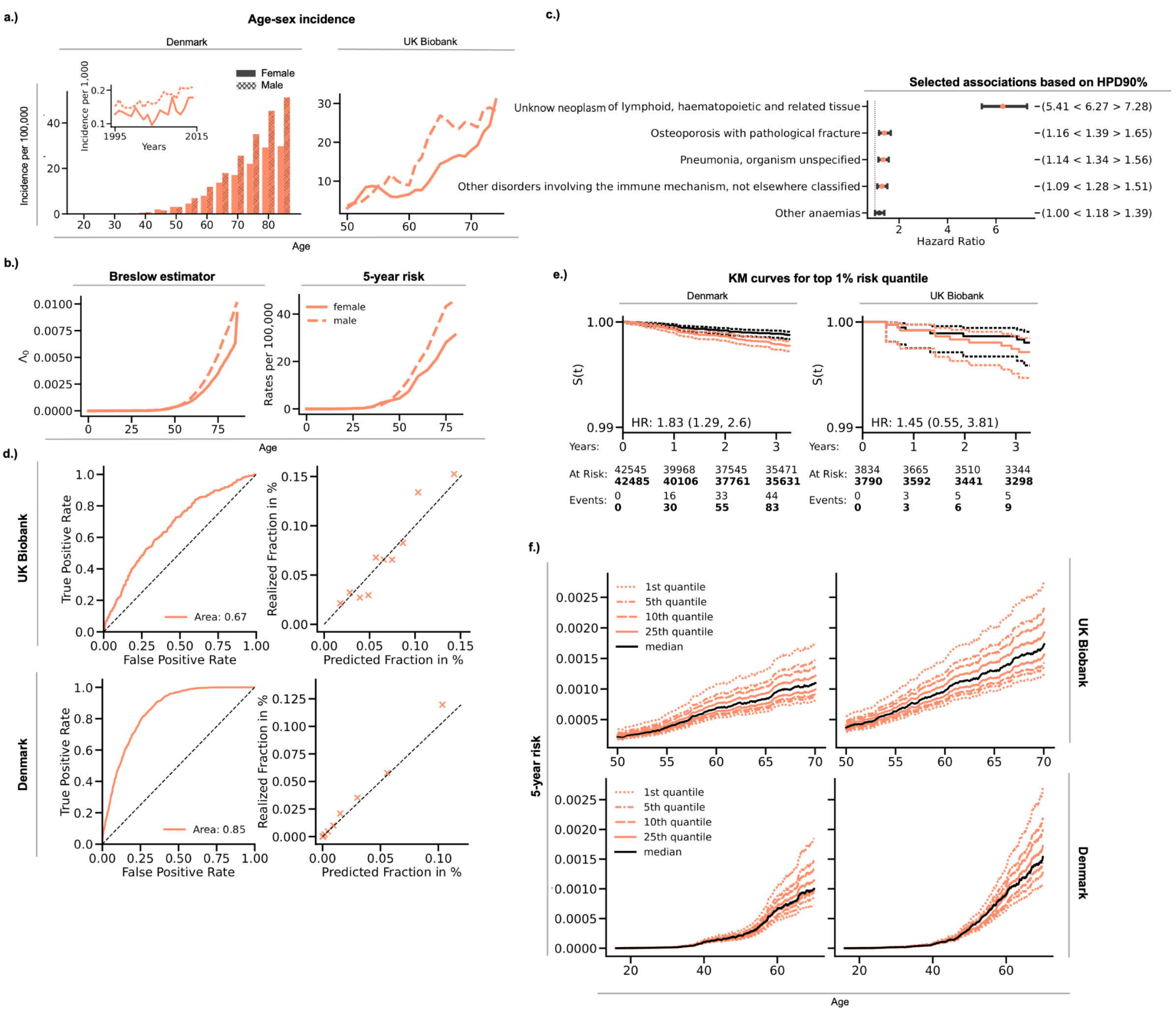

#### **SFC20:** AML

**AML**

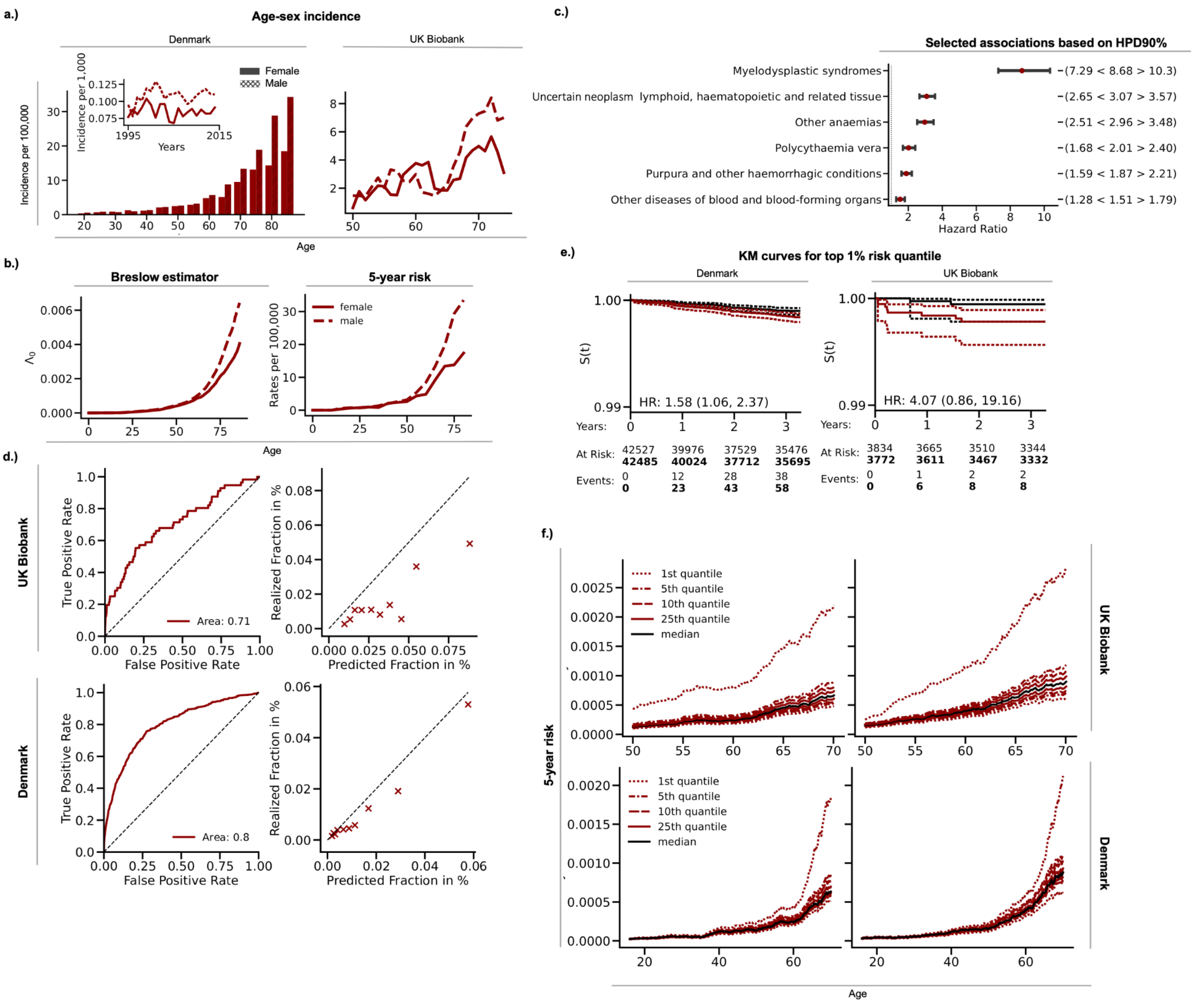
